# SARS-CoV-2 dynamics in New York City during March 2020–August 2023

**DOI:** 10.1101/2024.07.19.24310728

**Authors:** Wan Yang, Hilary Parton, Wenhui Li, Elizabeth A. Watts, Ellen Lee, Haokun Yuan

**Affiliations:** Department of Epidemiology, Columbia University, New York, NY, USA; New York City Department of Health and Mental Hygiene, Queens, NY, USA; Centers for Disease Control and Prevention, Atlanta, GA, USA

## Abstract

The severe acute respiratory syndrome coronavirus 2 (SARS-CoV-2) has been widespread since 2020 and will likely continue to cause substantial recurring epidemics. However, understanding the underlying infection burden (i.e., including undetected asymptomatic/mild infections) and dynamics, particularly since late 2021 when the Omicron variant emerged, is challenging due to the potential for asymptomatic and repeat SARS-CoV-2 infection, changes in testing practices, and changes in disease reporting. Here, we leverage extensive surveillance data available in New York City (NYC) and a comprehensive model-inference system to reconstruct SARS-CoV-2 dynamics therein from the pandemic onset in March 2020 to August 2023, and further validate the estimates using independent wastewater surveillance data. The validated model-inference estimates indicate a very high infection burden totaling twice the population size (>5 times documented case count) but decreasing infection-fatality risk (a >10-fold reduction) during the first 3.5 years. The detailed estimates also reveal highly complex variant dynamics and immune landscape, changing virus transmissibility, and higher infection risk during winter in NYC over this time period. These transmission dynamics and drivers, albeit based on data in NYC, may be relevant to other populations and inform future planning to help mitigate the public health burden of SARS-CoV-2.

## INTRODUCTION

The severe acute respiratory syndrome coronavirus 2 (SARS-CoV-2) emerged in late 2019. Within months, it quickly spread worldwide, prompting the World Health Organization (WHO) to declare coronavirus disease 2019 (COVID-19) a global public health emergency on January 30, 2020, a designation lasting for 3+ years through May 5, 2023 (*1*). Populations worldwide have experienced multiple COVID-19 pandemic waves, and will likely continue to endure recurring epidemics, even after the declared ending of the pandemic phase. Given the disease’s historical importance, high potential to cause future epidemics, and long-term health impacts (e.g., long-COVID (*2*)), it is important to better understand its transmission dynamics, infection burden, and severity over time.

Many studies have reported SARS-CoV-2 transmission dynamics during the initial and subsequent pandemic waves (*3*). However, transmission dynamics after the Omicron BA.1 wave remain less characterized. Many Omicron subvariants have emerged after BA.1, causing outbreaks with varying magnitude and quickly supplanting one another (*4*). While surveillance systems (e.g., registries of laboratory-reported cases and death certificates) can provide invaluable information, potential biases (e.g., due to differential test-seeking behaviors) could limit the understanding of epidemic dynamics (*5–7*). For example, underlying SARS-CoV-2 infection rates were not completely captured by surveillance based on clinical testing due to high rates of asymptomatic and mild infection (*8, 9*) and use of at-home testing, the results of which are not reported to health departments (*10*), nor were they captured by serologic surveys due to high rates of reinfection. Due to these limitations, to what extent populations are (re)infected by each subvariant and what drives the Omicron-subvariant waves – e.g., increased transmissibility and/or immune evasion – remain unclear.

In this study, we leverage extensive surveillance data available in New York City (NYC) and a comprehensive model-inference system to reconstruct the underlying SARS-CoV-2 transmission dynamics therein during March 2020 – August 2023. NYC is a densely populated, large, urban center with 8+ million people that became one of the first pandemic epicenters in March 2020. We have previously reported model-inference estimates for the first two pandemic waves (*11–13*). Here, we fit a more detailed model to age- and neighborhood-specific data of COVID-19 cases, emergency department (ED) visits, and deaths, and validate model-inference estimates using independent SARS-CoV-2 wastewater viral load data, i.e., measurements of population-level SARS-CoV-2 fecal shedding that are less subject to testing biases. The validated model-inference estimates allow quantification of weekly infection rates by each (sub)variant and key epidemiologic features including the underlying population susceptibility, variant-specific transmissibility, and infection-fatality risk (IFR) over time since the pandemic onset. Overall, we estimate a very high infection burden totaling twice the population size (>5 times the case count) but decreasing IFRs (a >10-fold reduction across all age groups), and highlight several key factors driving transmission dynamics, during the initial 3.5 years of SARS-CoV-2 circulation.

## RESULTS

### The model-inference system reconstructed underlying SARS-CoV-2 infection dynamics that are consistent with independent SARS-CoV-2 wastewater surveillance data

The model-inference system is able to recreate the epidemic curves of weekly cases, ED visits, and deaths, combining all ages (Fig 1A) and for individual age groups (Fig S1). Given the large uncertainty due to changes in clinical testing and reporting requirements, we further validate the estimates using independent SARS-CoV-2 wastewater surveillance data not used for model inference. As shown in Fig 1B, the estimated number of infectious people per 100,000 population per week closely tracked the measured SARS-CoV-2 load in wastewater. This close agreement is evident for all three major periods, i.e., the 2^nd^ wave mostly due to the ancestral and Iota variants during Fall 2020/Winter 2021 (Fig 1B, 1^st^ panel; Pearson correlation coefficient *r* = 0.91, 95% confidence interval [CI]: 0.84-0.95), the Delta wave during Summer/Fall 2021 (Fig 1B, 2^nd^ panel; *r* = 0.64, 95% CI: 0.29-0.84), and the Omicron period since late November 2021 (Fig 1B, 3^rd^ panel and inset for recent months; *r* = 0.89, 95% CI: 0.84-0.93). These results indicate the model-inference system adequately accounted for changing infection-detection rates over time, and accurately reconstructed the underlying SARS-CoV-2 infection rates and transmission dynamics during the study period.

**Fig 1.**
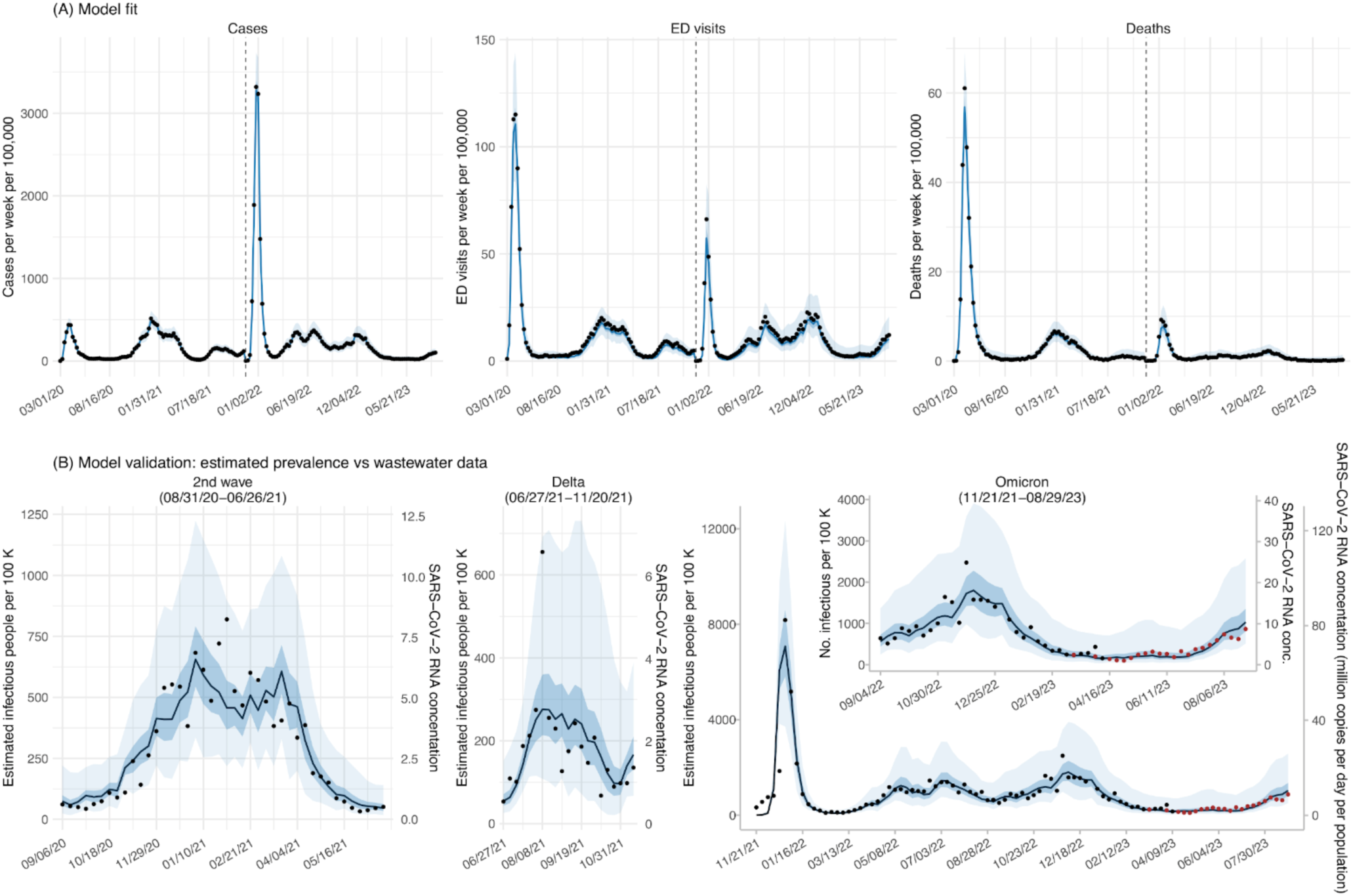
Model fit and validation. (A) Model fit to weekly number of COVID-19 cases, emergency department (ED) visits, and deaths, during the week starting 03/01/20 (mm/dd/yy) to the week starting 08/27/23 (see x-axis). Blue lines show the median estimates and blue areas show 50% (darker) and 95% (lighter) credible intervals (CrIs); dots show the corresponding observations. (B) Model validation using wastewater surveillance data, for the 2^nd^ wave (left panel), Delta wave (middle panel), the Omicron period (right panel). Lines and shaded areas show the estimated infection prevalence (i.e., the number of all infectious individuals including those not detected as cases; median, 50% and 95% CrIs; left y-axis). Dots show measured SARS-CoV-2 concentrations in wastewater (right y-axis, in million copies per day per population) for the corresponding weeks (black dots show measurements using RT-qPCR and red dots show measurements using RT-dPCR but converted to RT-qPCR equivalents; note that the wastewater concentrations are scaled for each wave/period to facilitate comparison with model estimates; see Methods for details).

### Overview of the COVID-19 pandemic/epidemic dynamics through August 2023

During the study period (the week starting March 1, 2020, to the week starting August 27, 2023), 3.2 million confirmed and probable cases were reported to the NYC Department of Health and Mental Hygiene (NYC Health Department) (or 38.2% of the size of the city’s population; Table 1). However, estimated infections totaled 17.4 million [95% credible interval (CrI): 14.2-21.5], more than five times the documented case count. During the pre-Omicron period (March 2020 – November 2021), the model-inference system estimated cumulative infections totaling 53.7% (95% CrI: 43.7-64.6%) of the size of the city’s population; these estimates include all infections, and do not distinguish between initial and subsequent infections for the same individual. Most of these infections were caused by the ancestral and Iota variants during the 1^st^ and 2^nd^ waves (estimated 38.0% of the size of the city’s population, 95% CrI: 19.7-75.4%), followed by Delta (8.4%, 95% CrI: 4.9-21.3%) and Alpha (2.9%, 95% CrI: 1.5-5.6%).

**Table 1.** Estimated cumulative infection rate, by variant. For ease of comparison, we convert the number of detected cases and estimated infections to percentage relative to the size of NYC’s population (columns labeled “% population”), i.e., this percentage does not refer to unique individuals detected as cases or estimated to experience infections.

| Variant | Main circulation period | Cases detected |  | Estimated infections |  |
| --- | --- | --- | --- | --- | --- |
|  |  | n (×1000)<sup>a</sup> | % population<sup>b</sup> | n (×1000)<sup>a</sup> | % population<sup>b</sup> |
| All |  | 3,207 | 38.2% | 17,383 (14,154, 21,549) | 207% (168.5%, 256.6%) |
| Pre-Omicron |  | 1,162 | 13.8% | 4,509 (3,673, 5,423) | 53.7% (43.7%, 64.6%) |
| Omicron |  | 2,046 | 24.4% | 12,875 (10,481, 16,126) | 153.3% (124.8%, 192%) |
| BA.1 | 12/12/2021 - 02/05/2022 | 1,024 | 12% | 3,468 (2,188, 5,624) | 41.3% (26.1%, 67%) |
| Ancestral/Iota | 03/08/2020 - 04/17/2021 | 834 | 9.9% | 3,195 (1,653, 6,334) | 38% (19.7%, 75.4%) |
| BA.5 | 06/05/2022 - 12/03/2022 | 289 | 3.4% | 2,107 (1,148, 4,589) | 25.1% (13.7%, 54.6%) |
| XBB.1.5 | 11/27/2022 - 08/12/2023 | 175 | 2.1% | 1,783 (790, 5,018) | 21.2% (9.4%, 59.7%) |
| BA.2.12.1 | 04/10/2022 - 07/16/2022 | 145 | 1.7% | 881 (462, 1,948) | 10.5% (5.5%, 23.2%) |
| Delta | 06/27/2021 - 12/11/2021 | 225 | 2.7% | 704 (327, 1,792) | 8.4% (3.9%, 21.3%) |
| BA.2 | 02/13/2022 - 07/23/2022 | 128 | 1.5% | 621 (332, 1,375) | 7.4% (4%, 16.4%) |
| BQ.1 | 10/02/2022 - 01/07/2023 | 61 | 0.73% | 591 (311, 1,338) | 7% (3.7%, 15.9%) |
| BQ.1.1 | 10/09/2022 - 01/28/2023 | 58 | 0.7% | 566 (295, 1,287) | 6.7% (3.5%, 15.3%) |
| XBB | 10/23/2022 - 09/02/2023 | 37 | 0.44% | 439 (214, 1,181) | 5.2% (2.5%, 14.1%) |
| FL.1 | 05/07/2023 - 09/02/2023 | 13 | 0.16% | 243 (117, 709) | 2.9% (1.4%, 8.4%) |
| Alpha | 01/17/2021 - 06/12/2021 | 77 | 0.92% | 240 (128, 471) | 2.9% (1.5%, 5.6%) |
| BA.4.6 | 06/19/2022 - 11/19/2022 | 32 | 0.39% | 238 (130, 523) | 2.8% (1.5%, 6.2%) |
| XBB.1.16 | 04/23/2023 - 09/02/2023 | 13 | 0.16% | 234 (100, 802) | 2.8% (1.2%, 9.5%) |
| BA.4 | 05/22/2022 - 09/24/2022 | 32 | 0.39% | 223 (121, 481) | 2.7% (1.4%, 5.7%) |
| EG.5 | 05/21/2023 - 09/02/2023 | 11 | 0.14% | 209 (99, 617) | 2.5% (1.2%, 7.3%) |
| BA.2.75 | 09/04/2022 - 08/19/2023 | 13 | 0.15% | 124 (62, 310) | 1.5% (0.7%, 3.7%) |
| XBB.1.9 | 03/05/2023 - 09/02/2023 | 5 | 0.056% | 75 (30, 268) | 0.9% (0.4%, 3.2%) |
| BF.7 | 08/07/2022 - 12/24/2022 | 8 | 0.1% | 74 (38, 172) | 0.9% (0.5%, 2%) |
| Epsilon | 12/27/2020 - 04/17/2021 | 10 | 0.11% | 25 (15, 45) | 0.3% (0.2%, 0.5%) |
| Gamma | 03/21/2021 - 07/17/2021 | 4 | 0.052% | 16 (8, 38) | 0.2% (0.1%, 0.5%) |
| Mu | 04/11/2021 - 09/25/2021 | 3 | 0.035% | 12 (6, 33) | 0.1% (0.1%, 0.4%) |

<sup>a</sup>The numbers are shown in thousands (i.e., ×1000). <sup>b</sup>The percentages are relative to the city’s population size of 8.39 million residents.

During the Omicron period (November 2021 – August 2023), (re)infections by the Omicron subvariants alone tripled, totaling 12.9 million (95% CrI: 10.5-16.1), or 153.3% (95% CrI: 124.8-192.0%) of the size of the city’s population. The BA.1 wave was the largest Omicron-subvariant wave thus far, infecting around 40% of the size of the city’s population, or 3.5 million people (95% CrI: 2.2-5.6), within roughly two months (Table 1 and Fig 2A). After BA.1 subsided, multiple Omicron subvariants circulated in NYC. By the end of August 2023, at least 14 Omicron subvariants including BA.1 had an estimated cumulative infection rate surpassing 1% of the city’s population size (vs. only four such variants prior to Omicron; Table 1). Multiple smaller Omicron-subvariant waves occurred, often with several subvariants cocirculating (Fig 2A). Most notably, the BA.2/BA.2.12.1 wave occurred during Spring/Summer 2022, the BA.5 wave during Summer/Fall 2022, and the XBB.1.5 wave during Winter 2023, each infecting around 20% of the size of the city’s population (Table 1 and Fig 2A).

**Fig 2.**
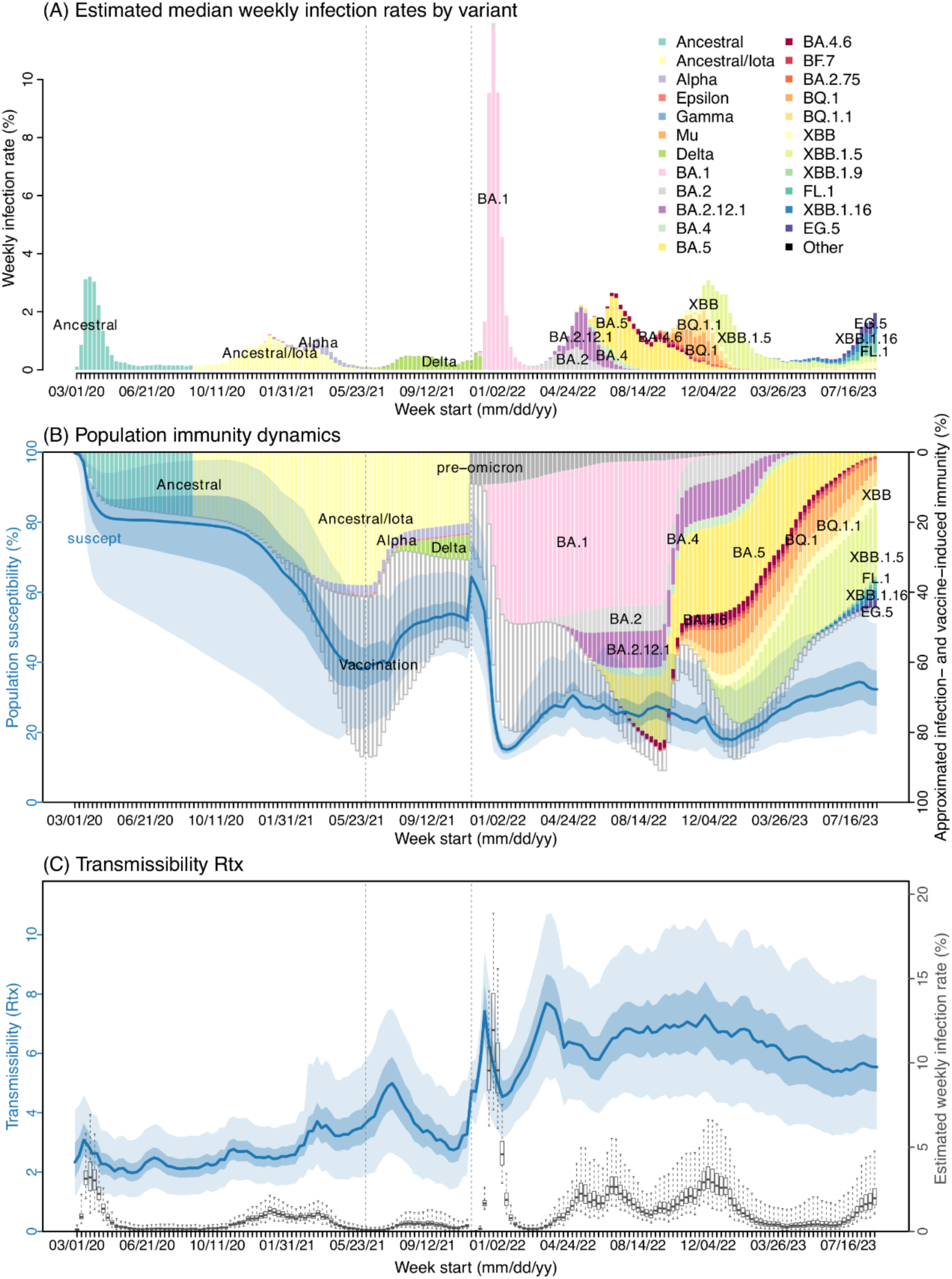
Estimated infection rates (A), population immunity dynamics (B), and virus transmissibility In (A), colored bars show estimated median weekly infection rates, for each variant (see legends). In (B), we overlay estimated population susceptibility [left y-axis; blue line = median, blue areas = 50% (darker) and 95% (lighter) CrIs], and proxies of cumulative infection (colored stacked bars from top to bottom, right y-axis; same legends as in A for different variants) and vaccine-induced immunity against infection (open bars; see Methods). In (C), we show estimated virus transmissibility [left y-axis; blue line = median, blue areas = 50% (darker) and 95% (lighter) CrIs] and infection rates [boxplot and right y-axis; middle bar = median, edges = 50% CrIs, and whiskers = 95% CrIs] for the corresponding weeks.

### Key factors driving SARS-CoV-2 transmission dynamics

SARS-CoV-2 transmission dynamics have been driven by multiple factors, including use of nonpharmaceutical interventions (NPIs), population immunity (due to prior infection and/or vaccination), new variants, and seasonal risk of infection (*11–13*), all of which were accounted by the model-inference system (see Methods). Since NPIs have become less prevalent during more recent waves (e.g., mask mandate in NYC schools was lifted in March 2022 (*14*)), here we focus on reporting the impact of the other aforementioned factors.

First, population susceptibility varies following surges in infections, vaccinations, and circulations of immune evasive variants (Fig 2B), and in turn determines the epidemic trajectory. Before the Delta wave, mixed immunity from both prior infections and vaccinations collectively lowered population susceptibility such that the sums (stacked bars, from top to bottom, in Fig 2B; see details in Methods) closely tracked the complement of estimated susceptibility (i.e., the estimated composite population immunity against infection; see blue line and shaded area in Fig 2B). The Delta variant partially evaded both infection- and vaccine-induced immunity (*15, 16*) such that the estimated susceptibility substantially increased during the Delta wave; the estimated population immunity was lower than the sum of prior infections and vaccinations (note this sum would roughly reflect the maximum of expected population immunity, should there be no immune evasion; see the stacked bars dipping below the blue line in Fig 2B). Nonetheless, strong mixed immunity at the time (>50%; see stacked bars and blue lines in Fig 2B) likely helped to temper the intensity of the Delta wave in Summer 2021.

Omicron BA.1 was highly immune evasive against all pre-existing variants (*17–19*). After adjusting for the lower vaccine effectiveness and weaker immunity from pre-Omicron infections (see Methods), the combined mixed immunity (stacked bars) closely matched the complement of estimated susceptibility (blue line) during the BA.1 wave (Fig 2B). It is evident from Fig 2B that rapid accumulation of BA.1 infection (pink bars) along with fast uptake of the 3^rd^ vaccine dose (open bars) at the time substantially increased population immunity, which likely accelerated the decline of BA.1. The large BA.1-infection-induced immunity also appeared to curb immediate surge of subsequent Omicron subvariants – particularly, BA.2/BA.2.12.1 and BA.5, even though these subvariants were able to partially evade that prior immunity (*20, 21*) (Fig 2B, see the stacked bars dipping below the blue line during Summer 2022). However, in Fall 2022/Winter 2023, infection-induced immunity appeared to come from a large number of post-BA.1 subvariants, accumulated through their continued spread (see increasing number of colors, each representing one subvariant, during the last part of the study period in Fig 2 A and B).

Second, virus transmissibility (*R_TX_*) can increase, helping newer variants to outcompete pre-existing ones. Here, to capture virus-specific transmissibility (*17, 22*), we separated the effects of changing population susceptibility, NPIs, and seasonal risk of infection. Unlike the effective reproduction number *R_t_* (i.e., the average number of secondary infections (*23*)), which can fluctuate due to the aforementioned effects, changes in *R_TX_* closely followed the surge of major variants (see large drops in *R_t_* around the pandemic onset due to NPIs in Fig S2A vs. the relative stable *R_TX_* in Fig 2C). That is, here *R_TX_* is akin to the basic reproduction number *R_0_* (i.e., the average number of secondary infections *in a naïve population* (*23*)) that measures the inherent transmissibility of a virus, and can be tracked over time for, e.g., new variants (vs. *R_0_* being estimated only at the pandemic onset when the entire population is susceptible; see Methods and refs (*17, 22*)).

In the above context, we estimate that virus transmissibility (*R_TX_*) has increased by nearly 3-fold in three years, but has appeared to level off since the latter half of 2022 (Fig 2C). Consistent with previous estimates (*13, 24*), Iota and Alpha increased virus transmissibility, allowing them to outcompete the ancestral variant during the 2^nd^ wave. In NYC, *R_TX_* increased by ∼20% during the 2^nd^ wave largely due to the mixed circulation of Iota and Alpha (Table 2). The Delta variant further increased virus transmissibility by another ∼30% (or ∼60% compared to the ancestral variant; Table 2), which, along with its immune evasive ability, allowed it to spread during Summer and Fall 2021 despite the relatively high population immunity at the time (Fig 2B). The Omicron BA.1 subvariant further increased virus transmissibility. In NYC, average *R_TX_* during the BA.1 wave was 2.3 times the 1st (ancestral variant) wave and further increased by ∼20% post-BA.1. Importantly, *R_TX_* remained around the same level through August 2023 (Table 2 and Fig 2C), suggesting immune evasion and waning immunity (Fig S2D) have been stronger drivers of the subvariant turnover since Summer 2022.

**Table 2.** Estimated virus transmissibility (*R_TX_*) and overall infection-fatality risk (IFR) during each wave/period. Numbers show the median estimate (and 95% credible intervals). Note the calendar periods here were chosen based on the rough timing of pandemic waves (March – May 2020 for the 1^st^ wave, October 2020 – May 2021 for the 2^nd^ wave, July – November 2021 for the Delta wave, and December 2021 – August 2023 for Omicron subvariants; see Fig 1), matching with the weekly intervals (hence the listed start dates) and excluding weeks with mixed circulation of variants (hence the missing weeks) in order to obtain more variant-specific estimates.

| Wave/variants | Calendar period | $R_{TX}$ | IFR (%) |
| --- | --- | --- | --- |
| 1st wave (ancestral) | 03/01/20 - 05/30/20 | 2.4 (1.33-3.68) | 1.429 (0.979-1.903) |
| 2nd wave<br>(ancestral/Iota/Alpha) | 10/04/20 - 05/29/21 | 2.86 (1.73-4.29) | 0.397 (0.246-0.6) |
| Delta | 07/04/21 - 12/04/21 | 3.75 (2.12-5.75) | 0.241 (0.094-0.602) |
| Omicron BA.1 | 12/12/21 - 02/26/22 | 5.46 (3.73-7.35) | 0.095 (0.047-0.166) |
| After BA.1 | 03/06/22 - 09/02/23 | 6.38 (4.15-9.37) | 0.056 (0.022-0.136) |

Third, seasonal conditions such as humidity and temperature may modulate the transmission of respiratory viruses including SARS-CoV-2 (*25–28*); in particular, low humidity and low temperature conditions commonly seen during the winter are conducive for SARS-CoV-2 survival (*25*). In addition, indoor crowding with reduced ventilation may also facilitate transmission (*29*). While infection rates could surge during summer months when new variants emerge, higher infection rates have occurred during winter months, peaking in December or January in NYC during the 3.5-year study period (Fig 3A). This pattern is further evident from Fig 3B, where the scaled infection rates during winter months were often more than twice as high as the summer months. This timing, despite multiple concurrent drivers including new variant emergence, highlights higher SARS-CoV-2 infection risk during the winter months in NYC during this study period.

**Fig 3.**
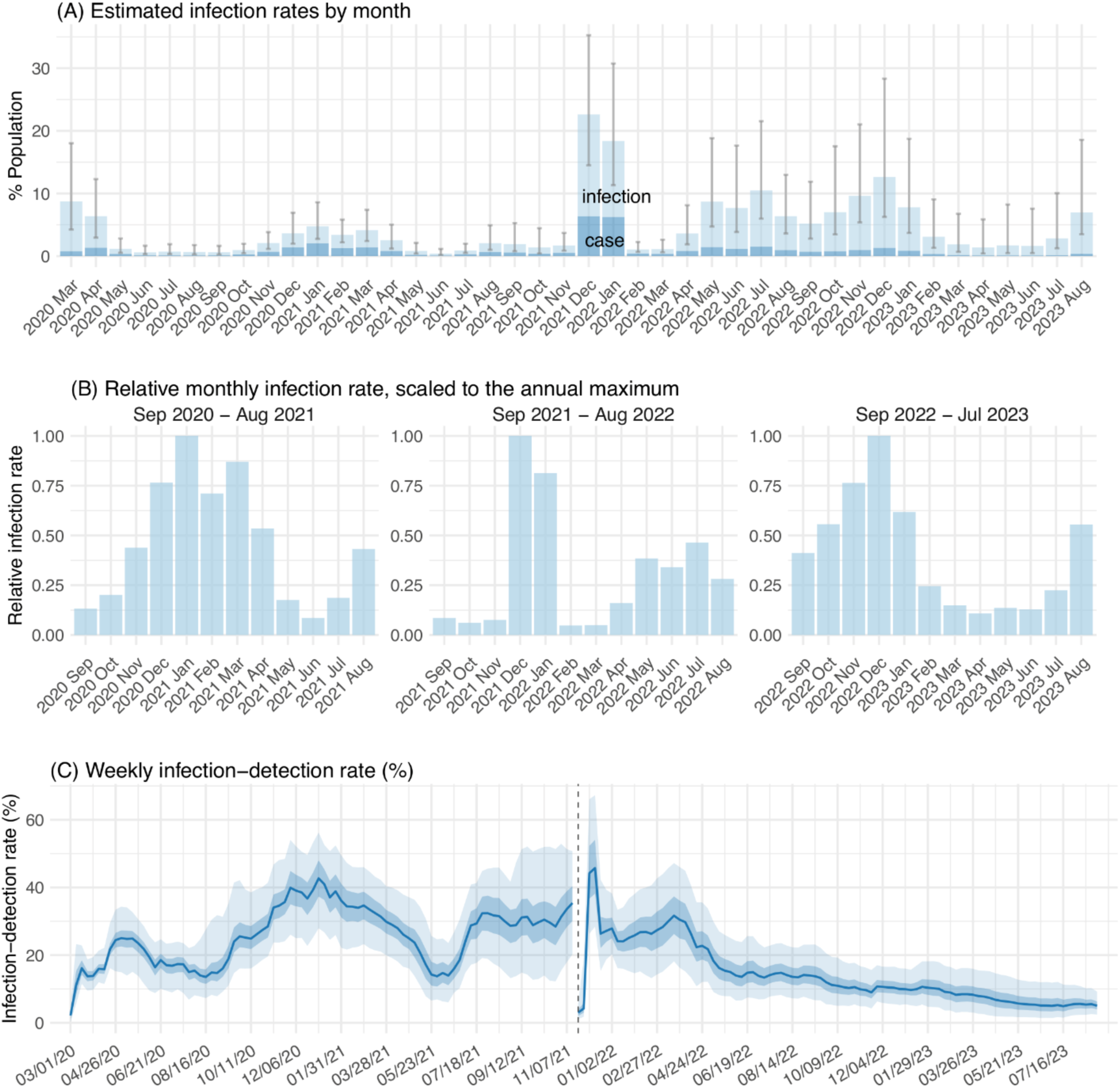
Infection annual pattern and detection rates. (A) shows estimated infection rates (light blue bars, full height; i.e., not stacked) and reported case rates (darker blue portion) by month; error bars show estimated 95% CrIs. To examine the infection annual pattern, (B) shows the monthly infection rates scaled to the annual maximum (here, a year starts in September, the start of fall/cold months in the Northern hemisphere, and ends in the next August, the end of winter/cold months in Southern hemisphere). March – August 2020 is not shown due to the incompleteness. (C) shows estimated infection-detection rate [blue line = median, blue areas = 50% (darker) and 95% (lighter) CrIs], for each week. Note the vertical dashed line indicates the week starting 11/21/21 when Omicron BA.1 was first detected in NYC, and estimates to the right of the dashed line are for Omicron (sub)variants alone.

### Changes in infection-detection rate

The infection-detection rate (i.e., case ascertainment rate) represents the proportion of infections detected as cases, and is crucial for accurate estimation of specific outcomes (e.g., infection rates and infection-fatality risk) to inform public health response (*30*). Estimating the infection-detection rate of SARS-CoV-2 has been challenging due to multiple factors (e.g., undetected asymptomatic/mild infections) (*5, 6, 30*). Here we resolved these challenges by comprehensive model inference (see Methods) and validated estimated infection-detection rates in NYC using independent wastewater surveillance data (Fig 1). In NYC, estimated infection-detection rates were very low at the onset of the 1^st^ pandemic wave and the Omicron BA.1 wave – only 2.1% (95% CrI: 0.2-4.5%) and 3.0% (95% CrI: 1.1-5.4%) of infections were detected as cases, respectively (Fig 3C). Estimated infection-detection rate increased substantially after the initial weeks of the pandemic but fluctuated over time (Fig 3C); the highest rates were estimated during the week of January 3, 2021 (42.7%, 95% CrI: 27.9-56.2%) before the Omicron variant emerged, and during the week of December 12, 2021 following the emergence of Omicron BA.1 (45.7%, 95% CrI: 28.4-67.2%). However, estimated infection-detection rate decreased steadily over time after Spring 2022 and was just ∼5% since Summer 2023 (Fig 3C), which is comparable to the initial weeks of the pandemic.

### Changes in infection-fatality risk (IFR)

IFR is a key indicator of COVID-19 severity. As reported previously, IFR of SARS-CoV-2 increased log-linearly with age (*31, 32*), particularly before mass-vaccination. Thus, we estimated IFR by age group (see Fig 4, Table 2 and Table S1). Consistent with previous reports (*12, 13, 33, 34*), estimated IFR in NYC was highest during the 1^st^ wave (March – May 2020). By the 2^nd^ wave (roughly October 2020 – May 2021), IFR had declined by more than half for most age groups, even though it increased transiently due to circulation of variants such as Iota and Alpha (Fig 4 D and E for those older than 65 years; and Table S2). During the latter half of 2021, IFR continued to decline, which likely was influenced by greater population immunity (Fig 2B).

**Fig 4.**
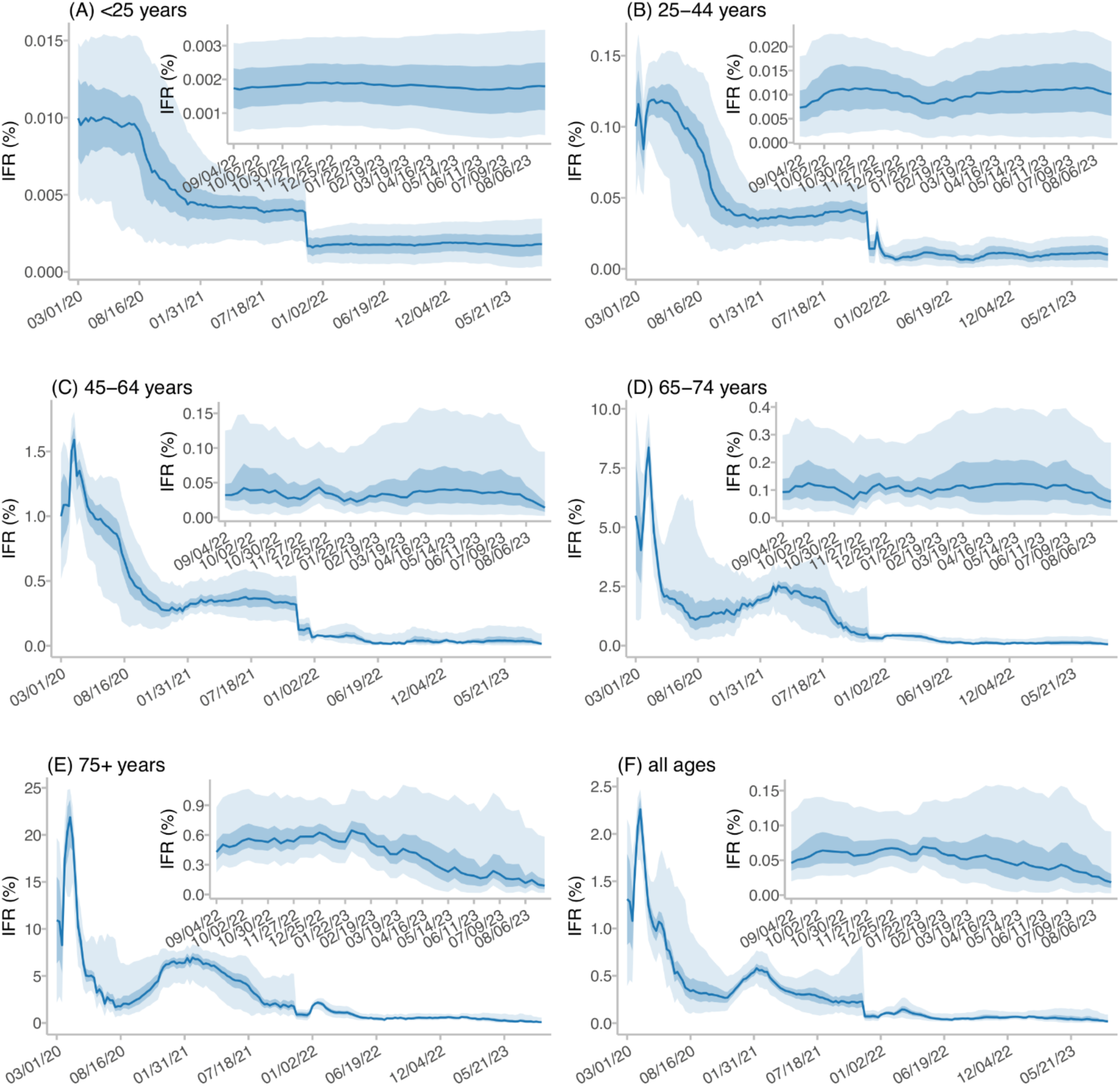
Estimated infection-fatality risk (IFR) over time, by age group (A-E) and overall (F). Blue lines and shaded areas show the median estimates and 50% (darker blue) and 95% (lighter blue) CrIs, for each week (see date of week start in mm/dd/yy in the x-axis). For clarity, insets show estimates during the most recent months.

Another substantial decline in IFR occurred following the circulation of Omicron BA.1, which was milder than pre-existing variants as reported previously (*35*). In NYC, estimated IFR during the Omicron BA.1 wave (December 2021-February 2022) declined by around half compared to the Delta wave (July – November 2021), for most age groups (Fig 4 and Table S1). After the Omicron BA.1 wave, overall IFR continued to decline, mostly driven by the lowering IFR among those aged 75 or older (Fig 4 E and F; Table 2 and Table S1). Starting the week of April 23, 2023, COVID-19 deaths were classified in NYC per a revised definition (see Methods). To examine the potential impact due to this change, we have also computed IFR including only weeks before the April 23, 2023 revision. The stratified IFR estimates were similar to those through the week of August 27, 2023 (Table S1).

## DISCUSSION

Using comprehensive model inference and data, we have reconstructed the transmission dynamics of SARS-CoV-2 in NYC during March 2020 – August 2023. The detailed model-inference estimates, further validated using independent SARS-CoV-2 wastewater surveillance data (Fig 1), can be used to inform future planning in the city (e.g., to gauge future SARS-CoV-2 infection burden and related health care needs). In addition, these estimates help to reveal the highly complex infection dynamics of SARS-CoV-2 and illustrate the key drivers in its continued spread, which may be shared by other populations. Below we focus on highlighting the general SARS-CoV-2 dynamics and key driving mechanisms.

By the end of August 2023, the estimated infection rate totaled twice the population size, indicating the majority of NYC residents may have had at least 2 (re)infections during the first 3.5 years. In addition, 81% of the population received the primary COVID-19 vaccine series, 40% have had an additional monovalent dose, and 23% have had either two additional monovalent doses or a bivalent vaccine (NYC vaccination data; as of 8/31/2023). In combination, these estimates and data suggest a high mixed population immunity. The high mixed population immunity would likely help mitigate the severity of future epidemics. Estimated IFR dropped by more than 10-fold for most age groups by August 2023, potentially attributable to multiple factors, including accumulated mixed immunity, access to improved treatment, and circulation of the milder Omicron subvariants. The potential long-term population health impact of this high infection rate is uncertain given the possibility of post-acute sequelae of SARS-CoV-2 infection (i.e., long-COVID (*2, 36*)).

As noted previously (*3*), the ability of SARS-CoV-2 to sustain continued spread in an already highly infected/vaccinated population has largely come from new variants, which can evade prior immunity and/or increase transmissibility. However, the dynamics and relative importance of these drivers have changed over time. Here, our estimates for NYC help to inform the interaction of these drivers during the first 3.5 years. When the underlying infection rate was relatively low (e.g., the first two waves), our estimates showed increased virus transmissibility predominantly drove SARS-CoV-2 variant dynamics (e.g., Alpha outcompeting pre-existing variants). As infections and immunity accumulated, we found stronger immune evasion allowed new variants to outcompete pre-existing and co-circulating subvariants, though transmissibility could also increase (Delta and BA.1 are both exemplars). By mid-2022, virus transmissibility appeared to stabilize after a nearly 3-fold increase (Fig 2C). Meanwhile, immune evasion continued but appeared to occur across multiple subvariants, each with a smaller subset of mutations, which may have allowed them to co-circulate and traverse pockets of resusceptible subpopulations (Fig 2B, see small changes in susceptibility after the Omicron BA.1 wave, despite substantial infections by 10+ Omicron subvariants). Whether this is a typical pathway of viral evolution to endemicity or whether another major Omicron BA.1-like new variant would emerge due to the nearly saturated immune landscape remains unknown.

The decrease in COVID-19 testing and data collection since early 2022 has raised concerns of timely situational awareness including new variant detection (*37–39*). Since late 2022/early 2023, the United States national surveillance strategy (*40, 41*) has further shifted to mainly monitoring infection trends and severity (e.g., hospitalizations and mortality), along with genomic surveillance and wastewater surveillance. Here, we estimated very low initial infection-detection rates – roughly, only 1 in 50 infections were detected – at the onset of the first pandemic wave and Omicron BA.1 wave in NYC. In addition, we found that population mobility (an indicator of community mitigation via social distancing (*11, 42–44*)) was inversely correlated with infection-detection rates during the initial weeks – that is, the lack of community mitigation coincided with low infection-detection rates at the time (see preliminary analysis in Table S2). The low infection-detection rates may have facilitated unchecked silent spread of SARS-CoV-2 during those initial weeks, as it likely did in other places (*45*). While the infection-detection rate increased by more than 10-fold during the pandemic, it has again declined to a very low level (Fig 3C). A fuller appreciation of under-detection in the design and implementation of surveillance systems is thus needed, as are innovative approaches to increase detection and awareness (e.g., wastewater surveillance with timely data sharing (*46*)).

Lastly, we note several study limitations. First, we did not account for population migration, which could lead to overestimation of the increase in susceptibility. In particular, the increase in population susceptibility after the Omicron BA.1 wave could be in part due to incoming population with a higher susceptibility than local residents (as NYC likely had a larger Omicron BA.1 wave and higher vaccination coverage than elsewhere), rather than entirely due to immune evasion of subsequent Omicron subvariants. Second, due to the discontinuation of SafeGraph mobility data (*47*), for weeks in 2023, we used mobility trends constructed based on historical data during the pandemic years 2020-2022 (vs. real-time mobility data for weeks before 2023; see Methods) to account for the impact of NPIs. However, we do not expect this to substantially affect the model-inference estimates, as the historical mobility trend was consistent with real-time subway ridership data (Fig S3). Third, the variant proportions among sequenced samples were used to estimate the variant-specific infection rates. However, as these samples may not be representative of the NYC population, estimates may reflect biases in the populations for which SARS-CoV-2 testing and sequencing were conducted. Lastly, per recommendation of the Council of State and Territorial Epidemiologists (CSTE), COVID-19-associated deaths were classified using a revised definition based solely on cause of death listed on the death certificate, for weeks from April 23, 2023 onwards. This revised definition could lead to missing COVID-19-associated deaths and thus underestimation of IFR afterwards. Nonetheless, a similar decline in IFR was estimated in the stratified analysis excluding weeks after the revision (Table S1), indicating a true continued decline in IFR after the Omicron BA.1 wave.

In summary, using comprehensive epidemiological data and model inference, we have described potential transmission dynamics of SARS-CoV-2 during its first 3.5 years of circulation in NYC, a large, urban center. Study findings highlight immune evasion, transmissibility increases, and higher infection risk during winter as key transmission drivers during the study period; these may be observed in other populations and could inform future planning to help mitigate the public health burden of SARS-CoV-2.

## METHODS

### Data sources and processing

For the model-inference system, we utilized multiple sources of epidemiologic data, including confirmed and probable COVID-19 cases, ED visits, deaths, vaccination, and variant proportions. As done and described previously (*11–13*), we aggregated all COVID-19 confirmed and probable cases (*48, 49*), COVID-19-associated ED visits (*13, 50*), and COVID-19-associated deaths (*49*) reported to the NYC Health Department by age group (<1, 1-4, 5-14, 15-24, 25-44, 45-64, 65-74, and 75+ year-olds), neighborhood of residence (42 United Hospital Fund neighborhoods in NYC (*51*)), and week of occurrence (*13*). For mortality, we note a change in COVID-19-associated death definitions. From March 1, 2020 – April 2, 2023, COVID-19-associated deaths included 1) deaths occurring in persons with laboratory-confirmed SARS-CoV-2 infection (i.e., confirmed COVID-19-associated death) at any point (March 1, 2020 – July 23, 2020), within 60 days (July 24, 2020 – August 2, 2021), or within 30 days (August 3, 2021 – April 2, 2023) of diagnosis; and 2) deaths with COVID-19, SARS-CoV-2 or a similar term listed on the death certificate as an immediate, underlying, or contributing cause of death but without laboratory-confirmation of COVID-19 (i.e., probable COVID-19-associated death) (*52*). From April 3, 2023 through the week of August 27, 2023 (i.e., end of this study), COVID-19-associated deaths included any death where the death certificate included COVID-19 or a common variation of COVID-19, SARS-CoV-2, coronavirus, etc. (*53*). For vaccinations, we included all available vaccine doses to date (i.e., 1^st^ to 5^th^ dose), and aggregated data for each vaccine dose to the same age/neighborhood strata, by date of vaccination (*54*).

To model the impact of NPIs, as done previously (*11–13*), we used mobility data from SafeGraph (*47*) to adjust SARS-CoV-2 transmission rate. Note, however, the model-inference system also included a parameter to capture the overall impacts of NPIs not limited to mobility reduction (e.g., additional interventions such as masking; see below). The SafeGraph data were aggregated to the neighborhood level by week without age stratification, and available from the week of March 1, 2020 to the week of December 19, 2022. For the week of December 26, 2022 to the week of August 27, 2023 (i.e., end of our study period), a comparison of historical SafeGraph data (i.e., weeks during March 2020 – December 2022, using the maximum mobility recorded for the corresponding week of year to account for seasonal changes) showed a close agreement with real-time subway ridership data (Fig S3). Thus, we used historical SafeGraph data for those weeks.

To compute the variant-specific estimates, we used reported weekly percentage of individual variants among sequenced samples (*55, 56*). Variant proportion data started from the week of December 27, 2020, and likely did not fully capture the share of Iota, a major variant that emerged around Fall 2020. Therefore, we combined the ancestral and Iota variants when computing the total number of cases or infections attributable to these variants.

For model validation, we used SARS-CoV-2 wastewater surveillance data, available from August 31, 2020 onward. Specifically, SARS-CoV-2 RNA concentrations were measured at each of the city’s 14 wastewater treatment plants, often twice per week, using quantitative reverse transcription polymerase chain reaction (RT-qPCR) assays during August 31, 2020 – April 11, 2023 and reverse transcription digital PCR (RT-dPCR) assays from November 1, 2022 through the week of August 27, 2023 (i.e., end of this study). For weeks after April 11, 2023 when the samples were measured using RT-dPCR alone, we converted the RT-dPCR measurements to RT-qPCR equivalents, by multiplying a simple conversion ratio (i.e., the mean of all RT-qPCR measurements dividing the mean of all RT-dPCR measurements during November 1, 2022 – April 11, 2023 when both assays were conducted). To compute the citywide weekly per-capita SARS-CoV-2 wastewater concentrations, we first averaged the per-capita SARS-CoV-2 concentrations (i.e., normalized by sewershed flow rate and population size) for each week and sewershed, and then further aggregated the sewershed-level measurements to the city level (i.e., weighted mean per the population size).

This activity was classified as public health surveillance and exempt from ethical review and informed consent by the Institutional Review Boards of both Columbia University and NYC Health Department.

### Model inference to estimate key epidemiological variables and parameters

We used a model-inference system to estimate epidemiological variables and parameters based on case, ED visit, and mortality data, accounting for NPIs, vaccinations, under-detection of infection, and seasonal changes. Built on an approach described in Yang et al. (*13*), here the model-inference system additionally tracks the number of vaccinated individuals and accounts for all vaccine doses as done in ref (*57*). Briefly, the model-inference system uses a metapopulation network SEIRSV (Susceptible-Exposed-Infectious-(re)Susceptible-Vaccination) model (Eq. 1) to simulate the transmission of SARS-CoV-2 by age group and neighborhood:

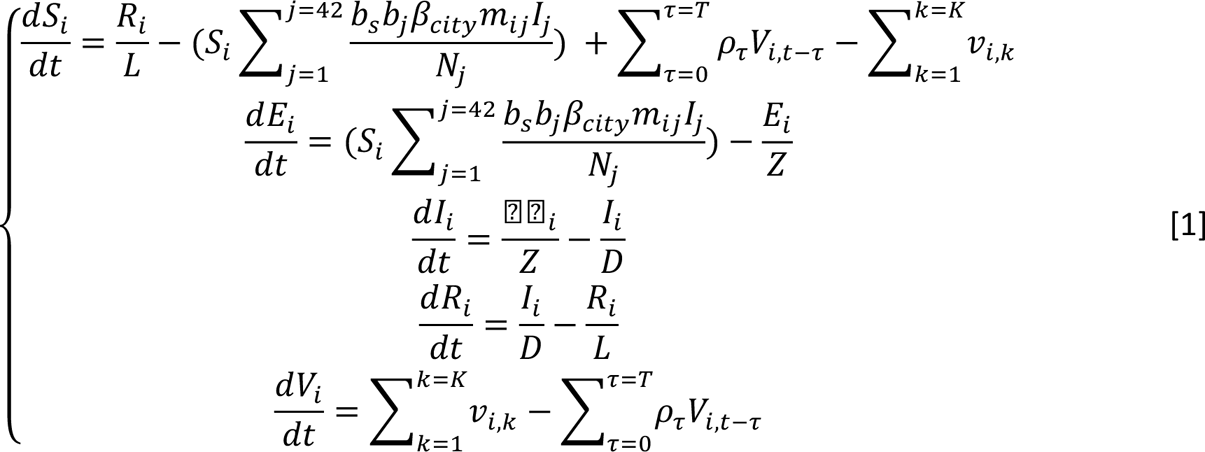

where *S_i_*, *E_i_*, *I_i_*, *R_i_*, *V_i_*, and *N_i_* are the number of susceptible, exposed (but not yet infectious), infectious, recovered and immune (i.e., protected against infection), vaccinated and immune individuals, and the total population (*58*), respectively, from a given age group (i.e., <1, 1-4, 5-14, 15-24, 25-44, 45-64, 65-74, or 75+ years) in neighborhood-*i* (*i* = 1,…42, for the 42 neighborhoods in the city). *β_city_* is the average citywide transmission rate; *b_s_* is the estimated seasonal trend (*12*). The term *b_i_* represents the neighborhood-level transmission rate relative to the city average. The term *m_ij_* represents the changes in contact rate in each neighborhood (for *i*=*j*) or spatial transmission from neighborhood-*j* to *i* (for *i*≠*j*) and was computed based on the mobility data (*12*). Here, we did not explicitly model the impact of individual NPI such as masking, due to the lack of data and the minor impact of masking at the population level (estimated 5-20% reduction (*11, 42, 59*)). Rather, to account for the overall impact of NPIs including masking, we scaled the mobility data by a multiplicative factor to capture the overall NPI effectiveness when computing *m_ij_* (*12*). *Z*, *D, and L* are the latency period, infectious period, and immunity period, respectively. Note that as all state variables and parameters are time varying and for each age group separately, Eq. 1 omits time (*t*) and age in the subscripts.

To account for vaccination, 𝜐*_i,k_*_,._ is the number of neighborhood-*i* residents who were immunized after the *k*-th dose (*k* = 1, 2, …, 5 here for up to 5 doses of vaccines to date) at the time step (*t*), and was computed using vaccination data adjusting for vaccine effectiveness (VE) against infection (*60–64*). Thus, the term 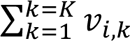_,._ represents the total number of neighborhood-*i* residents immunized by any dose of vaccine at the time step. The term 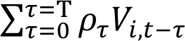 accounts for the waning of vaccine protection against infection, where *V_i,t-τ_* is the number of neighborhood-*i* residents who got vaccinated *τ* days ago and lost protection on day-*t*, and *ρ_τ_* is the VE waning probability computed based on VE duration data (*63*). Note, here we focused on modeling the impact of vaccination on population susceptibility, and that the posterior estimates of population susceptibility were made along with other factors (e.g., infection) using several data streams and model inference as described below.

Using the model-simulated number of infections occurring each day, we further computed the number of cases, ED visits, and deaths each week to match with the observations, as described in (*12, 13*). Using cases as an example, we multiplied the model-simulated number of new infections per day by the infection-detection rate (i.e., case ascertainment rate, or the fraction of infections reported as cases), and further distributed these estimates in time per a distribution of time-from-infection-to-case-detection (Table S3); we then aggregated the daily lagged, simulated estimates to weekly totals for model inference.

Each week, the system uses the ensemble adjustment Kalman filter (EAKF) (*65*) to compute the posterior estimates of model state variables and parameters based on the model (prior) estimates and observed case, ED visit, and mortality data per Bayes’ rule (*12, 13*). In particular, model posterior estimates include 1) the underlying infection rate including those not reported as cases, for each week (Fig 2A and C); 2) the number of susceptible individuals (i.e., *S_i_*), which provides estimates of population susceptibility over time (Fig 2B); 3) the citywide transmission rate (*β_city_*) and infectious period (see estimates in Fig S2 B and C), which we multiplicatively combined to compute the time-varying virus transmissibility (*R_TX_*, as measure of variant-specific infectiousness as described in (*17, 22*); Fig 2C); and 4) other key parameters such as the infection-detection rate (Fig 3C), IFR (Fig 4), and the real-time production number (*R_t_*; see estimates in Fig S2A).

We ran the model-inference system for the pre-Omicron and Omicron periods, separately. For the pre-Omicron period, we initiated the system at the week of March 1, 2020 (i.e., the week the first cases were detected in NYC), and ran it continuously through the week of December 5, 2021 (i.e., the week before the Omicron BA.1 variant was detected in >50% of sequenced cases). For the Omicron period, we reinitiated the system at the week of November 21, 2021 and ran it continuously through the end of the study period; given the initial overlap with the Delta variant in November/December 2021, we computed the number of cases, ED visits, and deaths due to Omicron based on the variant proportion data and used those variant-specific estimates for inference. To account for model uncertainty, we ran the model-inference system 10 times, each with 500 ensemble members randomly drawn from the initial prior ranges (Table S3), and combined the posteriors from all runs, as done in (*12*).

### Model validation using SARS-CoV-2 wastewater surveillance data

To validate model-inference estimates, we compared the infection prevalence estimates (i.e., the estimated number of infectious individuals, including those not detected as cases, in the population each week) to independent SARS-CoV-2 wastewater concentration data (i.e., the collective SARS-CoV-2 viral shedding of the population, regardless of clinical testing practices). While both quantities represent the presence of SARS-CoV-2 in the population, the measurements are on different scales and viral shedding per infection could vary by the infecting variant. Thus, for comparison, we separated the data into three periods: i) August 31, 2020 (i.e., the first day of wastewater surveillance) – June 26, 2021, predominantly the ancestral and Iota variants; ii) June 27, 2021 (i.e., the first week the share of Delta exceeding 50% among the sequenced samples) – November 20, 2021, predominantly the Delta variant; and iii) November 21, 2021 (i.e., the first week Omicron BA.1 was detected) – August 29, 2023 (i.e., the last wastewater sample during the study period), predominantly the Omicron subvariants. We scaled the wastewater measurements by multiplying the ratio of mean infection prevalence estimates and mean wastewater concentrations across all weeks of each period, and overlay the two time series for visual inspection (see Fig 1B).

### Estimating variant-specific infection rates

The weekly infection rate estimates from the model-inference system are based on surveillance data combining all reported variants and thus represent infections by any variant circulating during the week. To estimate the variant-specific infection rates for each week, we multiplied the overall infection rate estimate by the proportion among the sequenced samples for each variant during that week. To compute the total variant-specific infection rate, we then summed the weekly estimates across all weeks that a given variant was detected. For each variant, to identify the main circulation period (i.e., calendar weeks when 95% of all infections occurred), we recorded the first week that the cumulative infection rate surpassed 2.5% (i.e., the start) and 97.5% (i.e., the end) of the total.

### Qualitative illustration of immunity from vaccinations and infections by different variants

The model-inference system accounted for immunity conferred by prior infection and vaccination and waning (Eq. 1) to compute the posterior estimates of population susceptibility, using epidemiological data and the EAKF inference algorithm as described above. However, because the two immune components overlap (e.g., a recoveree could subsequently get vaccinated and have mixed immunity for both) and the EAKF may not perfectly preserve mass balance, it is difficult to separately quantify their contributions. Thus, to qualitatively examine the population immunity landscape, we used the rolling sum of prior infection as a proxy of infection-induced immunity and that of vaccinations as a proxy of vaccine-induced immunity (shown in Fig 2B). Specifically, the rolling sum of prior infection was computed by adding all estimated infections during the preceding 0.5*T_rs_* days (i.e., the estimated half time of immunity period; see Fig S2D). The rolling sum of vaccinations was computed by adding vaccinations of the primary series, 3^rd^, and 4^th^, 5^th^ dose during the preceding 0.5*T_vax_* days (*T_vax_* is the estimated vaccine-induced immunity period) and further multiplying the estimated variant-specific VE (Table S3).

## Data Availability

The SARS-CoV-2/COVID-19 cases, emergency department visits, mortality, and wastewater surveillance data were used with permission under a Data Use and Nondisclosure Agreement between the NYC DOHMH and Columbia University. The NYC DOHMH also has a comprehensive, publicly available data website here: https://github.com/nychealth/coronavirus-data. Additional data sources are detailed in the manuscript.

## Acknowledgments

This study was in part supported by the National Institute of Allergy and Infectious Diseases (AI145883 and AI175747), the Centers for Disease Control and Prevention (CDC) and the Council of State and Territorial Epidemiologists (CSTE; contract no.: NU38OT00297), and the National Science Foundation (DMS-2027369). The authors thank Lauren Firestein for overseeing the data use agreement and facilitating data sharing for this project; Ramona Lall for providing syndromic surveillance emergency department data; Iris Cheng for providing immunization data; Jubayer Ahmed, Nelson De La Cruz, Brandon Nguyen, and Greta Ohanian for managing and providing wastewater data; Elizabeth Luoma and Rebecca Rohrer for their management and provision of variant data; the NYC Health Department COVID data team for overarching data management and provision of data for this project; and Shama Ahuja, Sharon Greene, Scott Harper, Elizabeth Luoma, Aaron Olson, Enoma Omoregie, Mamta Parakh, Celia Quinn, Ulrike Siemetzki-Kapoor, Faten Taki, and Gretchen Van Wye for their input on this manuscript.

## Author contributions

WY designed the study, developed the model-inference system, performed the analysis, and wrote the first draft; HP and EL provided the COVID-19 case and emergency department visit data; WL provided the COVID-19-associated mortality data; EAW provided the SARS-CoV-2 wastewater surveillance data. HY compiled the mobility data. All authors contributed to the final draft.

## Supplemental Figures and Tables

**Fig S1.**
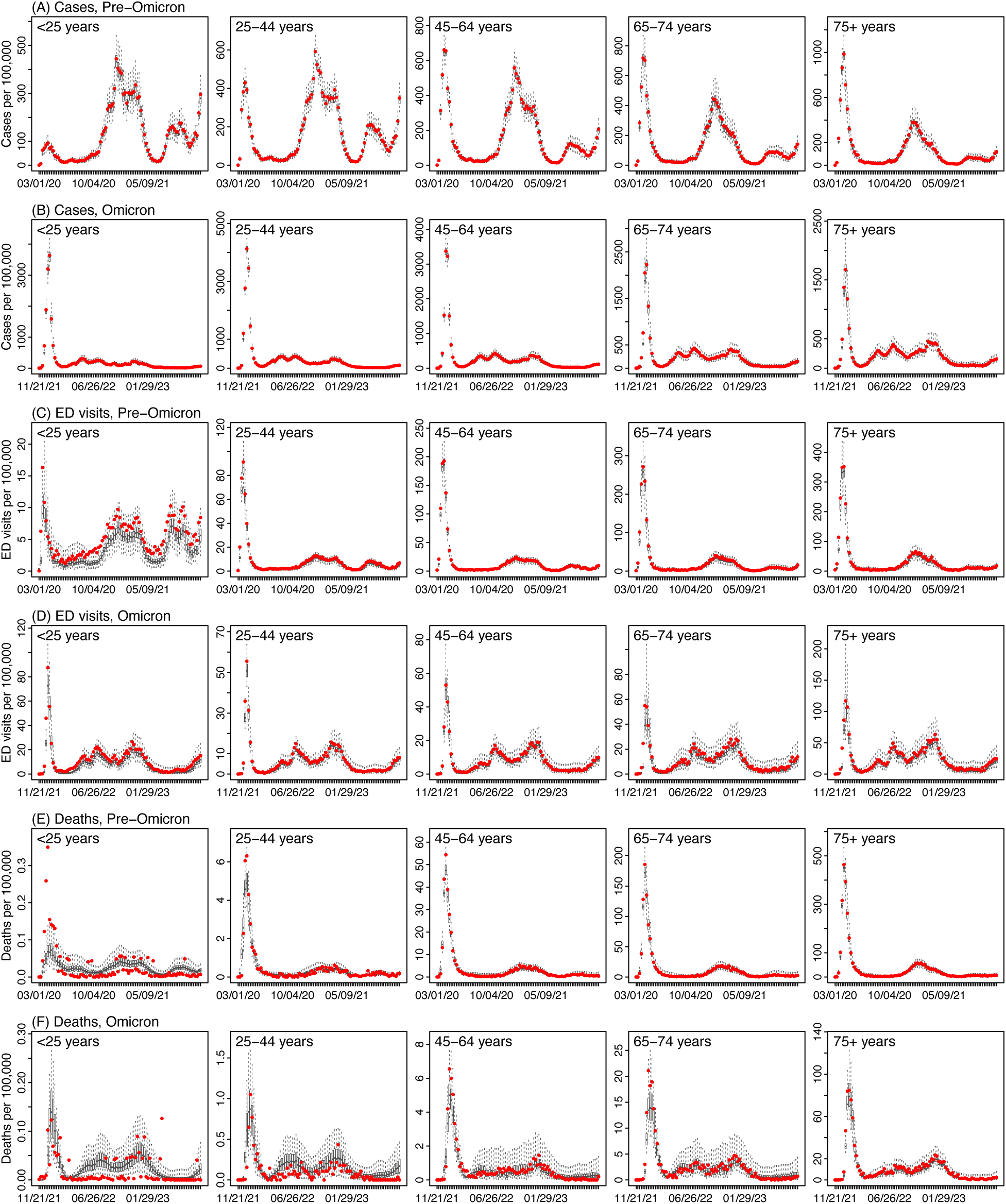
Model fits by age group. Boxplots show model estimates of COVID-19 cases (A and B), ED visits (C and D), and deaths (E and F) per 100,000 population (middle bar = median, edges = 50% CrIs, and whiskers = 95% CrIs), for each age group (see subtitle) and week (see x-axis, mm/dd/yy). Red dots show the corresponding observations.

**Fig S2.**
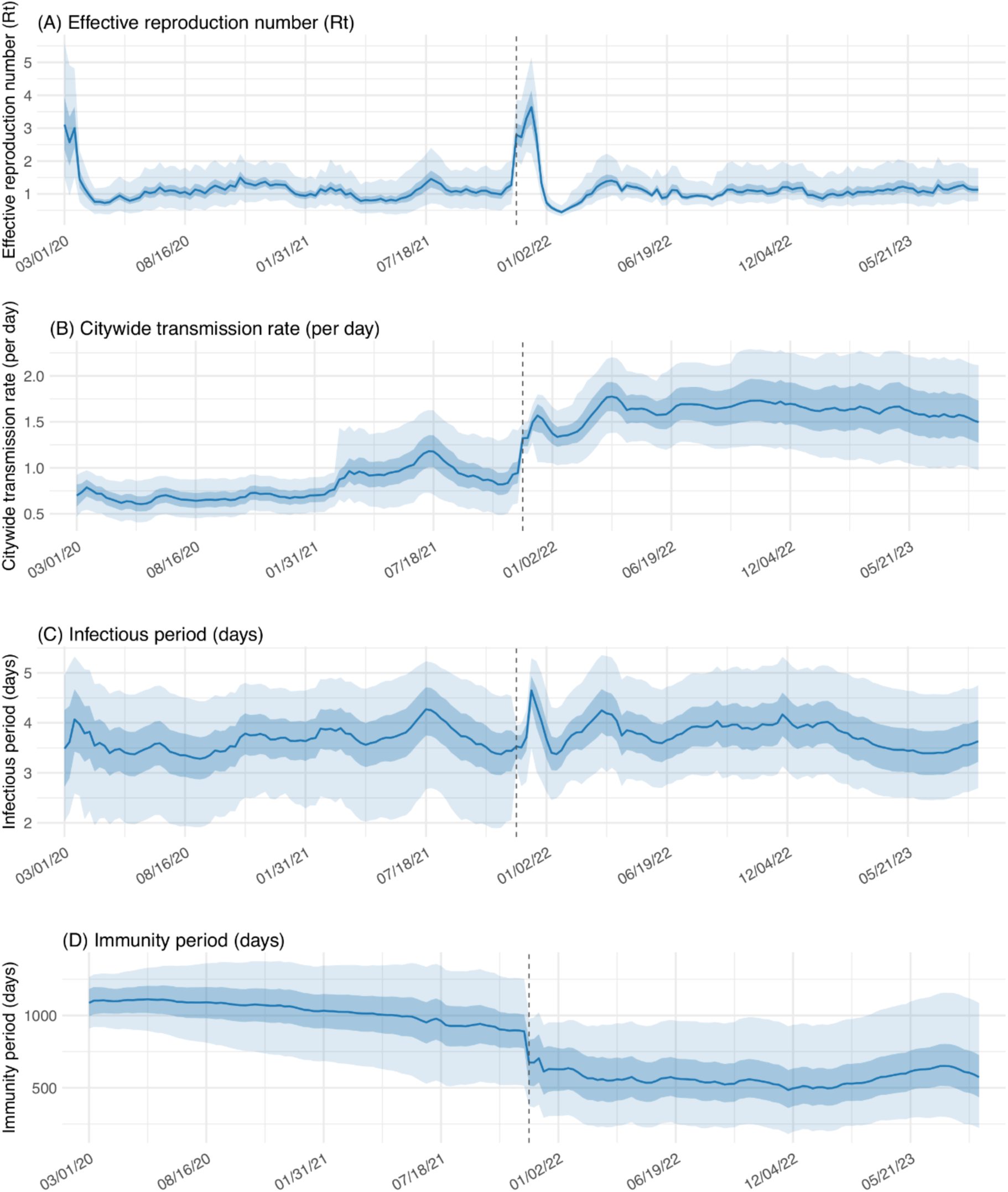
Estimates for key epidemiological variables: (A) the real-time effective reproduction number Rt, (B) citywide transmission rate (*β_city_* in Eq. 1) (C) infectious period, and (D) immunity period (*T_rs_* in Eq. 1). Blue lines and shaded areas show the median estimates and 50% (darker blue) and 95% (lighter blue) CrIs, for each week (see date of week start in mm/dd/yy in the x-axis). The vertical dashed line indicates the week starting 11/21/21 when Omicron BA.1 was first detected in NYC.

**Fig S3.**
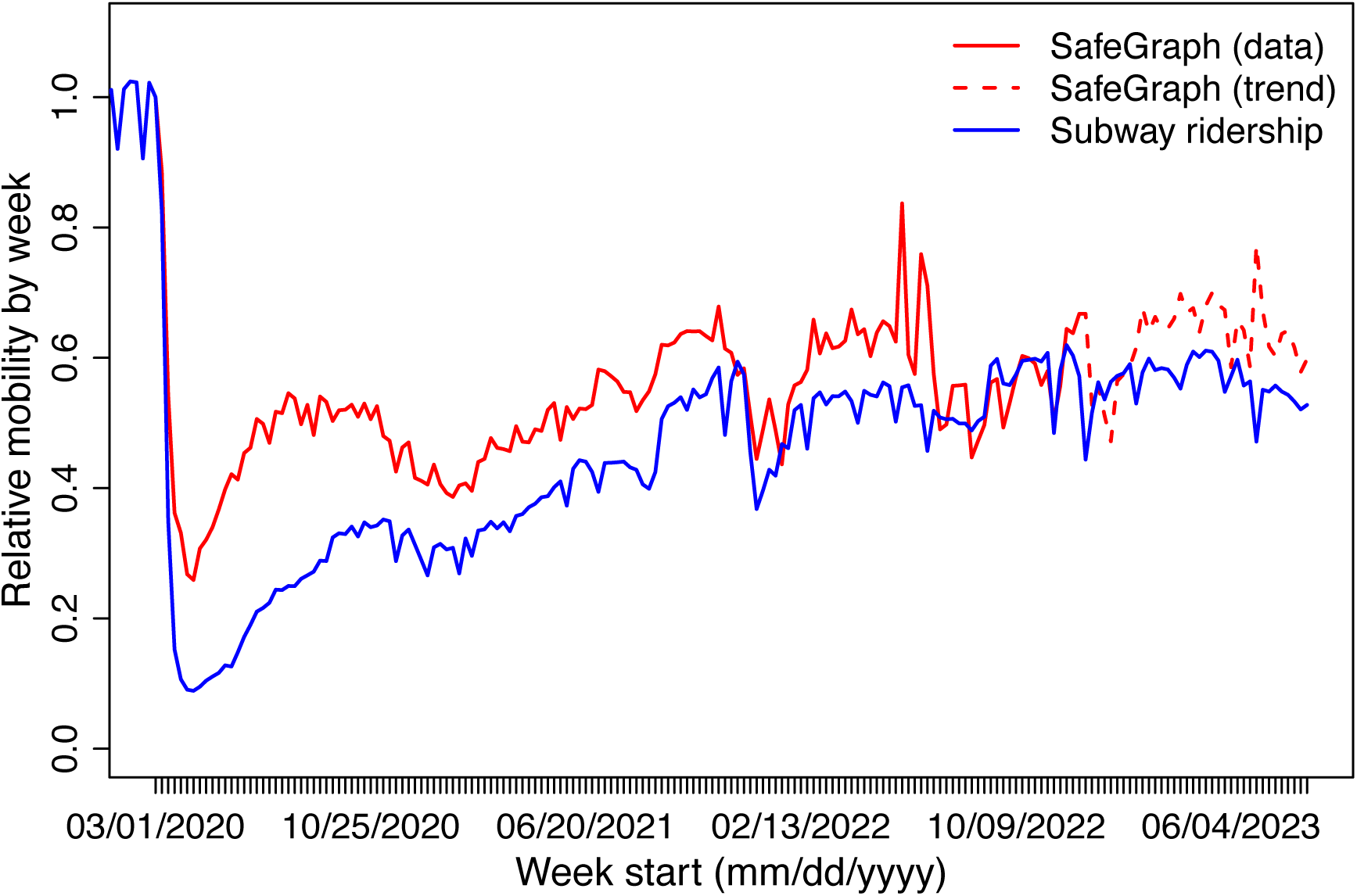
Trends in population mobility during the study period. Relative mobility for each week is computed by dividing the mobility for each week by that during the week starting March 1, 2020 (i.e., the pandemic onset in NYC). SafeGraph data were available from the week of March 1, 2020 to the week of December 19, 2022 (red solid line), and used in this study. For the week of December 26, 2022 to the week of August 27, 2023, we used trends constructed with historical SafeGraph data (i.e., weeks during March 2020 – December 2022, using the maximum mobility recorded for the corresponding week of year to account for seasonal changes; red dashed line). Real-time subway ridership data (blue line) are shown for comparison.

**Table S1.** Estimated infection-fatality risk (IFR, %), for individual age groups, during each wave/period. Numbers show the median estimate (and 95% credible intervals). The last two rows show stratified estimates for weeks post-BA.1, before and after the revision of COVID-19 death definition following the recommendation of the Council of State and Territorial Epidemiologists (CSTE).

| Wave/variants | Calendar period | all ages | <25 years | 25-44 years | 45-64 years | 65-74 years | 75+ years |
| --- | --- | --- | --- | --- | --- | --- | --- |
| 1st wave (ancestral) | 03/01/20 - 05/30/20 | 1.4291<br>(0.9793-1.903) | 0.0099<br>(0.0046-0.0151) | 0.1117<br>(0.0706-0.1466) | 1.1992<br>(0.8047-1.5056) | 4.6334<br>(2.7199-6.9133) | 12.2835<br>(7.3515-16.9023) |
| 2nd wave<br>(ancestral/Iota/Alpha) | 10/04/20 - 05/29/21 | 0.3965<br>(0.2461-0.6004) | 0.0047<br>(0.0019-0.0088) | 0.0379<br>(0.0174-0.0673) | 0.324<br>(0.2013-0.5231) | 1.9044<br>(0.9849-3.1934) | 5.5573 (3.238-7.5487) |
| Delta | 07/04/21 - 12/04/21 | 0.2406<br>(0.0936-0.6016) | 0.004 (0.0019-0.0061) | 0.04 (0.0209-0.0598) | 0.3436<br>(0.1906-0.5516) | 0.9821<br>(0.2883-2.6949) | 2.4551<br>(0.9768-5.4612) |
| Omicron BA.1 | 12/12/21 - 02/26/22 | 0.0945<br>(0.0474-0.1663) | 0.0017 (6e-04-0.0029) | 0.0106<br>(0.0042-0.0202) | 0.0842<br>(0.0441-0.147) | 0.3921<br>(0.2286-0.5353) | 1.5588<br>(0.9388-2.1269) |
| After BA.1 (all weeks) | 03/06/22 - 09/02/23 | 0.0561<br>(0.0219-0.1355) | 0.0018 (5e-04-0.0032) | 0.0098<br>(0.0016-0.0208) | 0.0352<br>(0.0071-0.1249) | 0.1457<br>(0.0471-0.3591) | 0.4927<br>(0.2496-1.0303) |
| After BA.1 (before revision of COVID-19 death definition)<sup>a</sup> | 03/06/22 - 04/22/23 | 0.0621<br>(0.0274-0.1349) | 0.0018 (5e-04-0.0032) | 0.0095<br>(0.0018-0.0202) | 0.0357<br>(0.0083-0.1207) | 0.1587<br>(0.0588-0.358) | 0.5885<br>(0.3186-1.0814) |
| After BA.1 (after revision of COVID-19 death definition)<sup>b</sup> | 04/23/23 - 09/02/23 | 0.0373<br>(0.0048-0.1375) | 0.0017 (3e-04-0.0034) | 0.0109<br>(0.0012-0.0227) | 0.0337<br>(0.0036-0.138) | 0.1055<br>(0.0107-0.3625) | 0.1951<br>(0.0355-0.8716) |

<sup>a</sup>Before April 22, 2023, COVID-19-associated deaths included deaths occurring in persons with laboratory-confirmed SARS-CoV-2 infection, and deaths with COVID-19, SARS-CoV-2, or a similar term listed on the death certificate as an immediate, underlying, or contributing cause of death but did not have laboratory-confirmation of COVID-19.
<sup>b</sup>From April 23, 2023 onward, COVID-19-associated deaths included any death where the death certificate includes COVID-19 or a common variation of COVID-19, SARS-CoV-2, coronavirus, etc.

**Table S2.** Correlation between infection-detection rate and population mobility, during the early weeks of the 1^st^ pandemic wave and the Omicron BA.1 wave. Numbers show the mean (and 95% confidence intervals [CI]) of Pearson’s correlation coefficients during the first 4, 6, and 8 weeks (column “First N weeks”) of the two pandemic waves. These results show estimated infection-detection rates were negatively correlated with observed population mobility during the early weeks of both pandemic waves; that is, initial increases in community mitigation via social distancing (as indicated by decreases in population mobility) coincided with increases in infection-detection rate. Note the Omicron BA.1 wave spread rapidly, peaked ∼1 month following the initial introduction, and subsided within ∼2 months; thus, the inverse association was the strongest during the first 4 weeks (vs. the first 8 weeks for the 1^st^ pandemic wave).

| Wave | First N weeks | Calendar period | Correlation (95% CI) | P-value |
| --- | --- | --- | --- | --- |
| 1<sup>st</sup> wave | 4 | 03/01/2020 - 04/04/2020 | -0.8 (-1, 0.69) | 0.2 |
|  | 6 | 03/01/2020 - 04/18/2020 | -0.82 (-0.98, -0.02) | 0.046 |
|  | 8 | 03/01/2020 - 05/02/2020 | -0.81 (-0.96, -0.24) | 0.015 |
| Omicron BA.1 wave | 4 | 11/21/2021 - 12/25/2021 | -0.96 (-1, -0.05) | 0.035 |
|  | 6 | 11/21/2021 - 01/08/2022 | -0.29 (-0.89, 0.68) | 0.58 |
|  | 8 | 11/21/2021 - 01/22/2022 | -0.28 (-0.82, 0.53) | 0.5 |

**Table S3.** Prior ranges for model parameters and variables.

| Period | Parameter | Symbol | Age group | Range | Note |
| --- | --- | --- | --- | --- | --- |
|  | Vaccine efficacy (VE) |  | All | Before Delta: VE1=85%, VE2 = 95%;<br>Delta: VE1 = 35.6%, VE2 = 88%, VE3 = 90%;<br>Omicron: VE1 = 35%, VE2 = 70%, VE3 = 70%, VE4 = 70%, VE5 (bivalent) = 90% | Based on VE against symptomatic disease |
| | VE waning | $\rho$ | All | $\rho(t) = 1/(1+\exp(-k * (t - t_{m.imm}/2))$;<br>for wildtype: $k = 0.026$; $t_{m.imm} = 322$;<br>for Delta: $k = 0.025$; $t_{m.imm} = 280$; for<br>Omicron: $k = 0.024$; $t_{m.imm} = 256$ | Parameter in the logistic function fitted based on data from UKHSA (ref 63) |
| Pre-Omicron | Citywide transmission rate (per day) | $\beta_{city}$ | <1 year | U [0.25, 0.5] | Uniform distribution, based on an overall transmission rate of 0.5–1 and adjusting for contact rate for the age group (same below) |
| Pre-Omicron | Citywide transmission rate (per day) | $\beta_{city}$ | 1-4 years | U [0.26, 0.52] | - |
| Pre-Omicron | Citywide transmission rate (per day) | $\beta_{city}$ | 5-14 years | U [0.33, 0.67] | - |
| Pre-Omicron | Citywide transmission rate (per day) | $\beta_{city}$ | 15-24 years | U [0.38, 0.77] | - |
| Pre-Omicron | Citywide transmission rate (per day) | $\beta_{city}$ | 25-44 years | U [0.69, 1.4] | - |
| Pre-Omicron | Citywide transmission rate (per day) | $\beta_{city}$ | 45-64 years | U [0.5, 1] | - |
| Pre-Omicron | Citywide transmission rate (per day) | $\beta_{city}$ | 65-74 years | U [0.25, 0.5] | - |
| Pre-Omicron | Citywide transmission rate (per day) | $\beta_{city}$ | 75+ years | U [0.25, 0.5] | - |
| Pre-Omicron | Latency period (days) | Z | All | U [2, 5] | - |
| Pre-Omicron | Infectious period (days) | D | All | U [2, 5] | - |
| Pre-Omicron | Immunity period (days) | L | <1 year | U [180, 550] | - |
| Pre-Omicron | Immunity period (days) | L | 1-4 years | U [910, 1300] | - |
| Pre-Omicron | Immunity period (days) | L | 5-14 years | U [910, 1300] | - |
| Pre-Omicron | Immunity period (days) | L | 15-24 years | U [910, 1300] | - |
| Pre-Omicron | Immunity period (days) | L | 25-44 years | U [910, 1300] | - |
| Pre-Omicron | Immunity period (days) | L | 45-64 years | U [910, 1300] | - |
| Pre-Omicron | Immunity period (days) | L | 65-74 years | U [910, 1300] | - |
| Pre-Omicron | Immunity period (days) | L | 75+ years | U [910, 1300] | - |
| Pre-Omicron | Time-to-detection, mean (days) | Td, mean | All | U [3, 8] | - |
| Pre-Omicron | Time-to-detection, SD (days) | Td,sd | All | U [1, 3] | - |
| Pre-Omicron | Infection-detection rate | IDR | <1 years | U [0.001, 0.05] | - |
| Pre-Omicron | Infection-detection rate | IDR | 1-4 years | U [1e-04, 0.05] | - |
| Pre-Omicron | Infection-detection rate | IDR | 5-14 years | U [1e-04, 0.05] | - |
| Pre-Omicron | Infection-detection rate | IDR | 15-24 years | U [1e-04, 0.05] | - |
| Pre-Omicron | Infection-detection rate | IDR | 25-44 years | U [0.001, 0.05] | - |
| Pre-Omicron | Infection-detection rate | IDR | 45-64 years | U [0.001, 0.05] | - |
| Pre-Omicron | Infection-detection rate | IDR | 65-74 years | U [0.001, 0.05] | - |
| Pre-Omicron | Infection-detection rate | IDR | 75+ years | U [0.001, 0.05] | - |
| Pre-Omicron | Infection-fatality risk | IFR | <1 years | U [5e-05, 0.00015] | - |
| Pre-Omicron | Infection-fatality risk | IFR | 1-4 years | U [5e-05, 0.00015] | - |
| Pre-Omicron | Infection-fatality risk | IFR | 5-14 years | U [5e-05, 0.00015] | - |
| Pre-Omicron | Infection-fatality risk | IFR | 15-24 years | U [5e-05, 0.00015] | - |
| Pre-Omicron | Infection-fatality risk | IFR | 25-44 years | U [5e-04, 0.0015] | - |
| Pre-Omicron | Infection-fatality risk | IFR | 45-64 years | U [0.005, 0.015] | - |
| Pre-Omicron | Infection-fatality risk | IFR | 65-74 years | U [0.01, 0.1] | - |
| Pre-Omicron | Infection-fatality risk | IFR | 75+ years | U [0.02, 0.2] | - |
| Pre-Omicron | Infection-to-ED ratio | EDR | <1 years | U [0.005, 0.1] | - |
| Pre-Omicron | Infection-to-ED ratio | EDR | 1-4 years | U [0.005, 0.1] | - |
| Pre-Omicron | Infection-to-ED ratio | EDR | 5-14 years | U [1e-04, 0.02] | - |
| Pre-Omicron | Infection-to-ED ratio | EDR | 15-24 years | U [0.001, 0.03] | - |
| Pre-Omicron | Infection-to-ED ratio | EDR | 25-44 years | U [0.001, 0.03] | - |
| Pre-Omicron | Infection-to-ED ratio | EDR | 45-64 years | U [0.001, 0.06] | - |
| Pre-Omicron | Infection-to-ED ratio | EDR | 65-74 years | U [0.01, 0.2] | - |
| Pre-Omicron | Infection-to-ED ratio | EDR | 75+ years | U [0.01, 0.3] | - |
| Omicron | Citywide transmission rate (per day) | $\beta_{city}$ | <1 years | U [0.61, 0.8] | - |
| Omicron | Citywide transmission rate (per day) | $\beta_{city}$ | 1-4 years | U [0.66, 0.85] | - |
| Omicron | Citywide transmission rate (per day) | $\beta_{city}$ | 5-14 years | U [0.91, 1.2] | - |
| Omicron | Citywide transmission rate (per day) | $\beta_{city}$ | 15-24 years | U [1, 1.3] | - |
| Omicron | Citywide transmission rate (per day) | $\beta_{city}$ | 25-44 years | U [1.6, 2.1] | - |
| Omicron | Citywide transmission rate (per day) | $\beta_{city}$ | 45-64 years | U [1.2, 1.6] | - |
| Omicron | Citywide transmission rate (per day) | $\beta_{city}$ | 65-74 years | U [0.6, 0.76] | - |
| Omicron | Citywide transmission rate (per day) | $\beta_{city}$ | 75+ years | U [0.58, 0.75] | - |
| Omicron | Latency period (days) | Z | <1 years | U [2.2, 3.8] | - |
| Omicron | Latency period (days) | Z | 1-4 years | U [2.1, 3.6] | - |
| Omicron | Latency period (days) | Z | 5-14 years | U [2, 3.5] | - |
| Omicron | Latency period (days) | Z | 15-24 years | U [2.1, 3.6] | - |
| Omicron | Latency period (days) | Z | 25-44 years | U [2.3, 3.8] | - |
| Omicron | Latency period (days) | Z | 45-64 years | U [2.2, 3.6] | - |
| Omicron | Latency period (days) | Z | 65-74 years | U [2.2, 3.7] | - |
| Omicron | Latency period (days) | Z | 75+ years | U [2.2, 3.7] | - |
| Omicron | Infectious period (days) | D | <1 years | U [2.2, 2.9] | - |
| Omicron | Infectious period (days) | D | 1-4 years | U [2.7, 3.5] | - |
| Omicron | Infectious period (days) | D | 5-14 years | U [2.9, 3.7] | - |
| Omicron | Infectious period (days) | D | 15-24 years | U [3.5, 4.5] | - |
| Omicron | Infectious period (days) | D | 25-44 years | U [3.2, 4.1] | - |
| Omicron | Infectious period (days) | D | 45-64 years | U [3.3, 4.2] | - |
| Omicron | Infectious period (days) | D | 65-74 years | U [2.5, 3.2] | - |
| Omicron | Infectious period (days) | D | 75+ years | U [2.7, 3.5] | - |
| Omicron | Immunity period (days) | L | <1 years | U [360, 470] | - |
| Omicron | Immunity period (days) | L | 1-4 years | U [360, 1000] | - |
| Omicron | Immunity period (days) | L | 5-14 years | U [360, 1100] | - |
| Omicron | Immunity period (days) | L | 15-24 years | U [360, 1000] | - |
| Omicron | Immunity period (days) | L | 25-44 years | U [360, 850] | - |
| Omicron | Immunity period (days) | L | 45-64 years | U [360, 1100] | - |
| Omicron | Immunity period (days) | L | 65-74 years | U [360, 990] | - |
| Omicron | Immunity period (days) | L | 75+ years | U [360, 960] | - |
| Omicron | Time-to-detection, mean (days) | Td, mean | <1 years | U [3.7, 5.2] | - |
| Omicron | Time-to-detection, mean (days) | Td, mean | 1-4 years | U [3.7, 5] | - |
| Omicron | Time-to-detection, mean (days) | Td, mean | 5-14 years | U [3.8, 4.9] | - |
| Omicron | Time-to-detection, mean (days) | Td, mean | 15-24 years | U [3.3, 4.3] | - |
| Omicron | Time-to-detection, mean (days) | Td, mean | 25-44 years | U [4, 4.9] | - |
| Omicron | Time-to-detection, mean (days) | Td, mean | 45-64 years | U [3.9, 4.8] | - |
| Omicron | Time-to-detection, mean (days) | Td, mean | 65-74 years | U [4.2, 5.2] | - |
| Omicron | Time-to-detection, mean (days) | Td, mean | 75+ years | U [4, 4.8] | - |
| Omicron | Time-to-detection, SD (days) | Td,sd | <1 years | U [1.5, 2.1] | - |
| Omicron | Time-to-detection, SD (days) | Td,sd | 1-4 years | U [1.6, 2.2] | - |
| Omicron | Time-to-detection, SD (days) | Td,sd | 5-14 years | U [1.7, 2.3] | - |
| Omicron | Time-to-detection, SD (days) | Td,sd | 15-24 years | U [1.6, 2.2] | - |
| Omicron | Time-to-detection, SD (days) | Td,sd | 25-44 years | U [1.9, 2.5] | - |
| Omicron | Time-to-detection, SD (days) | Td,sd | 45-64 years | U [2, 2.5] | - |
| Omicron | Time-to-detection, SD (days) | Td,sd | 65-74 years | U [1.7, 2.2] | - |
| Omicron | Time-to-detection, SD (days) | Td,sd | 75+ years | U [1.7, 2.1] | - |
| Omicron | Infection-detection rate | IDR | All | U [0.01, 0.05] | - |
| Omicron | Infection-fatality risk | IFR | <1 years | U [6.1e-06, 2.8e-05] | - |
| Omicron | Infection-fatality risk | IFR | 1-4 years | U [6.2e-06, 2.8e-05] | - |
| Omicron | Infection-fatality risk | IFR | 5-14 years | U [6.2e-06, 2.8e-05] | - |
| Omicron | Infection-fatality risk | IFR | 15-24 years | U [6e-06, 2.7e-05] | - |
| Omicron | Infection-fatality risk | IFR | 25-44 years | U [4.3e-05, 0.00024] | - |
| Omicron | Infection-fatality risk | IFR | 45-64 years | U [0.00039, 0.0021] | - |
| Omicron | Infection-fatality risk | IFR | 65-74 years | U [0.0015, 0.005] | - |
| Omicron | Infection-fatality risk | IFR | 75+ years | U [0.0037, 0.014] | - |
| Omicron | Infection-to-ED ratio | EDR | <1 years | U [0.0035, 0.029] | - |
| Omicron | Infection-to-ED ratio | EDR | 1-4 years | U [0.0019, 0.013] | - |
| Omicron | Infection-to-ED ratio | EDR | 5-14 years | U [0.00061, 0.0047] | - |
| Omicron | Infection-to-ED ratio | EDR | 15-24 years | U [0.001, 0.0083] | - |
| Omicron | Infection-to-ED ratio | EDR | 25-44 years | U [0.00097, 0.0069] | - |
| Omicron | Infection-to-ED ratio | EDR | 45-64 years | U [0.0028, 0.019] | - |
| Omicron | Infection-to-ED ratio | EDR | 65-74 years | U [0.0024, 0.017] | - |
| Omicron | Infection-to-ED ratio | EDR | 75+ years | U [0.006, 0.041] | - |

## References

1. E. Harris, WHO Declares End of COVID-19 Global Health Emergency. JAMA-J Am Med Assoc 329, (2023).

2. H. E. Davis, L. McCorkell, J. M. Vogel, E. J. Topol, Long COVID: major findings, mechanisms and recommendations. Nat Rev Microbiol 21, 133–146 (2023).

3. K. Koelle, M. A. Martin, R. Antia, B. Lopman, N. E. Dean, The changing epidemiology of SARS-CoV-2. Science 375, 1116–1121 (2022).

4. Global Initiative on Sharing All Influenza Data (GISAID), Tracking of hCoV-19 Variants. https://www.gisaid.org/hcov19-variants/

5. S. Riley, K. E. C. Ainslie, O. Eales, C. E. Walters, H. Wang, C. Atchison, C. Fronterre, P. J. Diggle, D. Ashby, C. A. Donnelly, G. Cooke, W. Barclay, H. Ward, A. Darzi, P. Elliott, Resurgence of SARS-CoV-2: Detection by community viral surveillance. Science 372, 990–995 (2021).

6. P. Elliott, M. Whitaker, D. Tang, O. Eales, N. Steyn, B. Bodinier, H. Wang, J. Elliott, C. Atchison, D. Ashby, W. Barclay, G. Taylor, A. Darzi, G. S. Cooke, H. Ward, C. A. Donnelly, S. Riley, M. Chadeau-Hyam, Design and Implementation of a National SARS-CoV-2 Monitoring Program in England: REACT-1 Study. Am J Public Health 113, 545–554 (2023).

7. J. Brainard, I. R. Lake, R. A. Morbey, N. R. Jones, A. J. Elliot, P. R. Hunter, Comparison of surveillance systems for monitoring COVID-19 in England: a retrospective observational study. Lancet Public Health 8, e850–e858 (2023).

8. P. Sah, M. C. Fitzpatrick, C. F. Zimmer, E. Abdollahi, L. Juden-Kelly, S. M. Moghadas, B. H. Singer, A. P. Galvani, Asymptomatic SARS-CoV-2 infection: A systematic review and meta-analysis. Proc Natl Acad Sci U S A 118, (2021).

9. Z. J. Feng, Q. Li, Y. P. Zhang, Z. Y. Wu, X. P. Dong, H. L. Ma, D. P. Yin, K. Lyu, D. Y. Wang, L. Zhou, R. Q. Ren, C. Li, Y. L. Wang, D. Ni, J. Zhao, B. Li, R. Wang, Y. Niu, X. H. Wang, L. J. Zhang, J. F. Sun, B. X. Liu, Z. Q. Deng, Z. T. Ma, Y. Yang, H. Liu, G. Shao, H. Li, Y. Liu, H. J. Zhang, S. Q. Qu, W. Lou, D. Shan, Y. H. Hu, L. Hou, Z. P. Zhao, J. M. Liu, H. Y. Wang, Y. J. Pang, Y. T. Han, Q. Y. Ma, Y. J. Ma, S. Chen, W. Li, R. T. Yang, Z. W. Li, Y. N. Guo, X. R. Liu, B. Jiangtulu, Z. X. Yin, J. Xu, S. Wang, L. Xiao, T. Xu, L. M. Wang, X. Qi, G. Q. Shi, W. X. Tu, X. M. Shi, X. M. Su, Z. J. Li, H. M. Luo, J. Q. Ma, J. M. McGoogan, The Epidemiological Characteristics of an Outbreak of 2019 Novel Coronavirus Diseases (COVID-19) - China, 2020. China CDC Weekly 2, 113–122 (2020).

10. B. Rader, A. Gertz, A. D. Iuliano, M. Gilmer, L. Wronski, C. M. Astley, K. Sewalk, T. J. Varrelman, J. Cohen, R. Parikh, Use of At-Home COVID-19 Tests—United States, August 23, 2021–March 12, 2022. Morbidity and Mortality Weekly Report 71, 489 (2022).

11. W. Yang, J. Shaff, J. Shaman, Effectiveness of non-pharmaceutical interventions to contain COVID-19: a case study of the 2020 spring pandemic wave in New York City. J R Soc Interface 18, 20200822 (2021).

12. W. Yang, S. Kandula, M. Huynh, S. K. Greene, G. Van Wye, W. Li, H. T. Chan, E. McGibbon, A. Yeung, D. Olson, A. Fine, J. Shaman, Estimating the infection-fatality risk of SARS-CoV-2 in New York City during the spring 2020 pandemic wave: a model-based analysis. The Lancet Infectious Diseases 21, 203–212 (2021).

13. W. Yang, S. K. Greene, E. R. Peterson, W. Li, R. Mathes, L. Graf, R. Lall, S. Hughes, J. Wang, A. Fine, Epidemiological characteristics of the B.1.526 SARS-CoV-2 variant. Science Advances 8, eabm0300 (2022).

14. The New York Times, New York City says it will end school mask and indoor proof-of-vaccination mandates. https://www.nytimes.com/2022/02/27/nyregion/new-york-mask-mandate-schools.html

15. P. Mlcochova, S. A. Kemp, M. S. Dhar, G. Papa, B. Meng, I. Ferreira, R. Datir, D. A. Collier, A. Albecka, S. Singh, R. Pandey, J. Brown, J. Zhou, N. Goonawardane, S. Mishra, C. Whittaker, T. Mellan, R. Marwal, M. Datta, S. Sengupta, K. Ponnusamy, V. S. Radhakrishnan, A. Abdullahi, O. Charles, P. Chattopadhyay, P. Devi, D. Caputo, T. Peacock, C. Wattal, N. Goel, A. Satwik, R. Vaishya, M. Agarwal, S.-C.-G. C. Indian, C. Genotype to Phenotype Japan, C.-N. B. C.-. Collaboration, A. Mavousian, J. H. Lee, J. Bassi, C. Silacci-Fegni, C. Saliba, D. Pinto, T. Irie, I. Yoshida, W. L. Hamilton, K. Sato, S. Bhatt, S. Flaxman, L. C. James, D. Corti, L. Piccoli, W. S. Barclay, P. Rakshit, A. Agrawal, R. K. Gupta, SARS-CoV-2 B.1.617.2 Delta variant replication and immune evasion. Nature 599, 114–119 (2021).

16. J. L. Bernal, N. Andrews, C. Gower, E. Gallagher, R. Simmons, S. Thelwall, J. Stowe, E. Tessier, N. Groves, G. Dabrera, R. Myers, C. N. J. Campbell, G. Amirthalingam, M. Edmunds, M. Zambon, K. E. Brown, S. Hopkins, M. Chand, M. Ramsay, Effectiveness of Covid-19 Vaccines against the B.1.617.2 (Delta) Variant. New England Journal of Medicine 385, 585–594 (2021).

17. W. Yang, J. L. Shaman, COVID-19 pandemic dynamics in South Africa and epidemiological characteristics of three variants of concern (Beta, Delta, and Omicron). Elife 11, e78933 (2022).

18. S. Cele, L. Jackson, D. S. Khoury, K. Khan, T. Moyo-Gwete, H. Tegally, J. E. San, D. Cromer, C. Scheepers, D. G. Amoako, F. Karim, M. Bernstein, G. Lustig, D. Archary, M. Smith, Y. Ganga, Z. Jule, K. Reedoy, S.-H. Hwa, J. Giandhari, J. M. Blackburn, B. I. Gosnell, S. S. Abdool Karim, W. Hanekom, M.-A. Davies, M. Hsiao, D. Martin, K. Mlisana, C. K. Wibmer, C. Williamson, D. York, R. Harrichandparsad, K. Herbst, P. Jeena, T. Khoza, H. Kløverpris, A. Leslie, R. Madansein, N. Magula, N. Manickchund, M. Marakalala, M. Mazibuko, M. Moshabela, N. Mthabela, K. Naidoo, Z. Ndhlovu, T. Ndung’u, N. Ngcobo, K. Nyamande, V. Patel, T. Smit, A. Steyn, E. Wong, A. von Gottberg, J. N. Bhiman, R. J. Lessells, M.-Y. S. Moosa, M. P. Davenport, T. de Oliveira, P. L. Moore, A. Sigal, Omicron extensively but incompletely escapes Pfizer BNT162b2 neutralization. Nature 602, 654–656 (2022).

19. J. R. C. Pulliam, C. van Schalkwyk, N. Govender, A. von Gottberg, C. Cohen, M. J. Groome, J. Dushoff, K. Mlisana, H. Moultrie, Increased risk of SARS-CoV-2 reinfection associated with emergence of Omicron in South Africa. Science 376, eabn4947 (2022).

20. Q. Wang, Y. Guo, S. Iketani, M. S. Nair, Z. Li, H. Mohri, M. Wang, J. Yu, A. D. Bowen, J. Y. Chang, J. G. Shah, N. Nguyen, Z. Chen, K. Meyers, M. T. Yin, M. E. Sobieszczyk, Z. Sheng, Y. Huang, L. Liu, D. D. Ho, Antibody evasion by SARS-CoV-2 Omicron subvariants BA.2.12.1, BA.4 and BA.5. Nature 608, 603–608 (2022).

21. Y. Cao, A. Yisimayi, F. Jian, W. Song, T. Xiao, L. Wang, S. Du, J. Wang, Q. Li, X. Chen, Y. Yu, P. Wang, Z. Zhang, P. Liu, R. An, X. Hao, Y. Wang, J. Wang, R. Feng, H. Sun, L. Zhao, W. Zhang, D. Zhao, J. Zheng, L. Yu, C. Li, N. Zhang, R. Wang, X. Niu, S. Yang, X. Song, Y. Chai, Y. Hu, Y. Shi, L. Zheng, Z. Li, Q. Gu, F. Shao, W. Huang, R. Jin, Z. Shen, Y. Wang, X. Wang, J. Xiao, X. S. Xie, BA.2.12.1, BA.4 and BA.5 escape antibodies elicited by Omicron infection. Nature 608, 593–602 (2022).

22. W. Yang, J. Shaman, Development of a model-inference system for estimating epidemiological characteristics of SARS-CoV-2 variants of concern. Nature Communications 12, 5573 (2021).

23. R. Anderson, C. Donnelly, D. Hollingsworth, M. Keeling, C. Vegvari, R. Baggaley, R. Maddren, Reproduction number (R) and growth rate (r) of the COVID-19 epidemic in the UK: methods of estimation, data sources, causes of heterogeneity, and use as a guide in policy formulation. The Royal Society 2020, (2020).

24. E. Volz, S. Mishra, M. Chand, J. C. Barrett, R. Johnson, L. Geidelberg, W. R. Hinsley, D. J. Laydon, G. Dabrera, A. O’Toole, R. Amato, M. Ragonnet-Cronin, I. Harrison, B. Jackson, C. V. Ariani, O. Boyd, N. J. Loman, J. T. McCrone, S. Goncalves, D. Jorgensen, R. Myers, V. Hill, D. K. Jackson, K. Gaythorpe, N. Groves, J. Sillitoe, D. P. Kwiatkowski, C.-G. U. consortium, S. Flaxman, O. Ratmann, S. Bhatt, S. Hopkins, A. Gandy, A. Rambaut, N. M. Ferguson, Assessing transmissibility of SARS-CoV-2 lineage B.1.1.7 in England. Nature 593, 266–269 (2021).

25. D. H. Morris, K. C. Yinda, A. Gamble, F. W. Rossine, Q. Huang, T. Bushmaker, R. J. Fischer, M. J. Matson, N. Van Doremalen, P. J. Vikesland, L. C. Marr, V. J. Munster, J. O. Lloyd-Smith, Mechanistic theory predicts the effects of temperature and humidity on inactivation of SARS-CoV-2 and other enveloped viruses. Elife 10, (2021).

26. L. C. Marr, J. W. Tang, J. Van Mullekom, S. S. Lakdawala, Mechanistic insights into the effect of humidity on airborne influenza virus survival, transmission and incidence. J R Soc Interface 16, (2019).

27. W. Yang, L. C. Marr, Mechanisms by which ambient humidity may affect viruses in aerosols. Appl Environ Microbiol 78, 6781–6788 (2012).

28. E. Huynh, A. Olinger, D. Woolley, R. K. Kohli, J. M. Choczynski, J. F. Davies, K. Lin, L. C. Marr, R. D. Davis, Evidence for a semisolid phase state of aerosols and droplets relevant to the airborne and surface survival of pathogens. Proc Natl Acad Sci U S A 119, (2022).

29. L. Morawska, J. W. Tang, W. Bahnfleth, P. M. Bluyssen, A. Boerstra, G. Buonanno, J. Cao, S. Dancer, A. Floto, F. Franchimon, C. Haworth, J. Hogeling, C. Isaxon, J. L. Jimenez, J. Kurnitski, Y. Li, M. Loomans, G. Marks, L. C. Marr, L. Mazzarella, A. K. Melikov, S. Miller, D. K. Milton, W. Nazaroff, P. V. Nielsen, C. Noakes, J. Peccia, X. Querol, C. Sekhar, O. Seppänen, S.-i. Tanabe, R. Tellier, K. W. Tham, P. Wargocki, A. Wierzbicka, M. Yao, How can airborne transmission of COVID-19 indoors be minimised? Environment International 142, 105832 (2020).

30. T. W. Russell, N. Golding, J. Hellewell, S. Abbott, L. Wright, C. A. B. Pearson, K. van Zandvoort, C. I. Jarvis, H. Gibbs, Y. Liu, R. M. Eggo, W. J. Edmunds, A. J. Kucharski, C. C.-w. group, Reconstructing the early global dynamics of under-ascertained COVID-19 cases and infections. BMC Med 18, 332 (2020).

31. A. T. Levin, W. P. Hanage, N. Owusu-Boaitey, K. B. Cochran, S. P. Walsh, G. Meyerowitz-Katz, Assessing the age specificity of infection fatality rates for COVID-19: systematic review, meta-analysis, and public policy implications. Eur J Epidemiol 35, 1123–1138 (2020).

32. M. O’Driscoll, G. R. Dos Santos, L. Wang, D. A. T. Cummings, A. S. Azman, J. Paireau, A. Fontanet, S. Cauchemez, H. Salje, Age-specific mortality and immunity patterns of SARS-CoV-2. Nature, (2020).

33. W. S. Hoogenboom, A. Pham, H. Anand, R. Fleysher, A. Buczek, S. Soby, P. Mirhaji, J. Yee, T. Q. Duong, Clinical characteristics of the first and second COVID-19 waves in the Bronx, New York: A retrospective cohort study. Lancet Reg Health Am 3, 100041 (2021).

34. N. G. Davies, C. I. Jarvis, C. C.-W. Group, W. J. Edmunds, N. P. Jewell, K. Diaz-Ordaz, R. H. Keogh, Increased mortality in community-tested cases of SARS-CoV-2 lineage B.1.1.7. Nature 593, 270–274 (2021).

35. N. Wolter, W. Jassat, S. Walaza, R. Welch, H. Moultrie, M. Groome, D. G. Amoako, J. Everatt, J. N. Bhiman, C. Scheepers, N. Tebeila, N. Chiwandire, M. du Plessis, N. Govender, A. Ismail, A. Glass, K. Mlisana, W. Stevens, F. K. Treurnicht, Z. Makatini, N.-y. Hsiao, R. Parboosing, J. Wadula, H. Hussey, M.-A. Davies, A. Boulle, A. von Gottberg, C. Cohen, Early assessment of the clinical severity of the SARS-CoV-2 omicron variant in South Africa: a data linkage study. The Lancet 399, 437–446 (2022).

36. National Academies of Sciences, Engineering, and Medicine, Long-Term Health Effects of COVID-19: Disability and Function Following SARS-CoV-2 Infection. P. A. Volberding, B. X. Chu, C. M. Spicer, Eds., (The National Academies Press, Washington, DC, 2024), pp. 264.

37. L. Ungar, Pandemic gets tougher to track as COVID testing plunges. https://apnews.com/article/covid-us-testing-decline-14bf5b0901260b063e4fa444633f4d31

38. M. Kekatos, COVID call centers and testing sites close in further sign US is moving past the pandemic. https://abcnews.go.com/Health/covid-call-centers-testing-sites-close-sign-us/story?id=97580639

39. R. Stein, As the pandemic ebbs, an influential COVID tracker shuts down. https://www.npr.org/sections/health-shots/2023/02/10/1155790201/as-the-pandemic-ebbs-an-influential-covid-tracker-shuts-down

40. B. J. Silk, H. M. Scobie, W. M. Duck, T. Palmer, F. B. Ahmad, A. M. Binder, J. A. Cisewski, S. Kroop, K. Soetebier, M. Park, A. Kite-Powell, A. Cool, E. Connelly, S. Dietz, A. E. Kirby, K. Hartnett, J. Johnston, D. Khan, S. Stokley, C. R. Paden, M. Sheppard, P. Sutton, H. Razzaghi, R. N. Anderson, N. Thornburg, S. Meyer, C. Womack, A. P. Weakland, M. McMorrow, L. R. Broeker, A. Winn, A. J. Hall, B. Jackson, B. E. Mahon, M. D. Ritchey, COVID-19 Surveillance After Expiration of the Public Health Emergency Declaration-United States, May 11, 2023. Mmwr-Morbid Mortal W 72, 523–528 (2023).

41. The Council of State and Territorial Epidemiologists, Association of Public Health Laboratories, Interim CSTE and APHL Strategic Framework for SARS-CoV-2 Infection and COVID-19 Surveillance: Priorities and Approaches for State, Territorial, Local, and Tribal Public Health Agencies. https://preparedness.cste.org/wp-content/uploads/2022/10/Interim-CSTE-APHL-COVID-Surveillance-Framework.pdf

42. W. Yang, J. Shaff, J. Shaman, Effectiveness of non-pharmaceutical interventions to contain COVID-19: a case study of the 2020 spring pandemic wave in New York City. J R Soc Interface 18, 20200822 (2021).

43. A. Lasry, D. Kidder, M. Hast, J. Poovey, G. Sunshine, K. Winglee, N. Zviedrite, F. Ahmed, K. A. Ethier, CDC Public Health Law Program, New York City Department of Health and Mental Hygiene, Louisiana Department of Health, Public Health Seattle and King County, San Francisco COVID-Response Team, Alameda County Public Health Department, San Mateo County Health Department, Marin County Division of Public Health, Timing of Community Mitigation and Changes in Reported COVID-19 and Community Mobility - Four U.S. Metropolitan Areas, February 26-April 1, 2020. MMWR. Morbidity and mortality weekly report 69, 451–457 (2020).

44. M. U. G. Kraemer, C. H. Yang, B. Gutierrez, C. H. Wu, B. Klein, D. M. Pigott, C.-D. W. G. Open, L. du Plessis, N. R. Faria, R. Li, W. P. Hanage, J. S. Brownstein, M. Layan, A. Vespignani, H. Tian, C. Dye, O. G. Pybus, S. V. Scarpino, The effect of human mobility and control measures on the COVID-19 epidemic in China. Science 368, 493–497 (2020).

45. R. Li, S. Pei, B. Chen, Y. Song, T. Zhang, W. Yang, J. Shaman, Substantial undocumented infection facilitates the rapid dissemination of novel coronavirus (SARS-CoV-2). Science 368, 489–493 (2020).

46. P. M. DeJonge, Wastewater surveillance data as a complement to emergency department visit data for tracking incidence of influenza A and respiratory syncytial virus—Wisconsin, August 2022–March 2023. MMWR. Morbidity and Mortality Weekly Report 72, (2023).

47. SafeGraph, Weekly Patterns: Foot Traffic Data To Understand The COVID-19 Pandemic. https://www.safegraph.com/weekly-foot-traffic-patterns

48. Centers for Disease Control and Prevention, National Notifiable Diseases Surveillance System (NNDSS) - Coronavirus Disease 2019 (COVID-19). https://ndc.services.cdc.gov/conditions/coronavirus-disease-2019-covid-19/

49. New York City Department of Health and Mental Hygiene, Defining confirmed and probable cases and deaths. https://www1.nyc.gov/site/doh/covid/covid-19-data.page

50. R. Lall, J. Abdelnabi, S. Ngai, H. B. Parton, K. Saunders, J. Sell, A. Wahnich, D. Weiss, R. W. Mathes, Advancing the Use of Emergency Department Syndromic Surveillance Data, New York City, 2012-2016. Public Health Rep 132, 23s–30s (2017).

51. NewYork City Department of Health and Mental Hygiene, NYC UHF 42 Neighborhoods. http://a816-dohbesp.nyc.gov/IndicatorPublic/EPHTPDF/uhf42.pdf

52. New York City Department of Health and Mental Hygiene (DOHMH) COVID-19 Response Team, Preliminary Estimate of Excess Mortality During the COVID-19 Outbreak — New York City, March 11–May 2, 2020. MMWR. Morbidity and mortality weekly report 69, 603–605 (2020).

53. New York City Department of Health and Mental Hygiene, NYC Coronavirus Disease 2019 (COVID-19) Data. 1/10/2024. https://github.com/nychealth/coronavirus-data

54. New York City Department of Health and Mental Hygiene, NYC Coronavirus 2019 (COVID-19) Vaccine Data. https://github.com/nychealth/covid-vaccine-data

55. C. N. Thompson, S. Hughes, S. Ngai, J. Baumgartner, J. C. Wang, E. McGibbon, K. Devinney, E. Luoma, D. Bertolino, C. Hwang, K. Kepler, C. Del Castillo, M. Hopkins, H. Lee, A. K. DeVito, J. L. Rakeman, PhD, A. D. Fine, Rapid Emergence and Epidemiologic Characteristics of the SARS-CoV-2 B.1.526 Variant - New York City, New York, January 1-April 5, 2021. MMWR Morb Mortal Wkly Rep 70, 712–716 (2021).

56. New york City Department of Health and Mental Hygiene, Variants. https://github.com/nychealth/coronavirus-data/tree/master/variants

57. W. Yang, J. Shaman, Development of Accurate Long-lead COVID-19 Forecast. PLoS Comput Biol 19, e1011278 (2023).

58. New York City Department of Health and Mental Hygiene, NYC DOHMH population estimates, modified from US Census Bureau interpolated intercensal population estimates, 2000-2018. Updated August 2019.

59. J. X. Benjamin, J Cowling, Escandón, Re: Effectiveness of public health measures in reducing the incidence of covid-19, SARS-CoV-2 transmission, and covid-19 mortality: systematic review and meta-analysis. BMJ, (2021. https://www.bmj.com/content/375/bmj-2021-068302/rr-14).

60. F. P. Polack, S. J. Thomas, N. Kitchin, J. Absalon, A. Gurtman, S. Lockhart, J. L. Perez, G. Perez Marc, E. D. Moreira, C. Zerbini, R. Bailey, K. A. Swanson, S. Roychoudhury, K. Koury, P. Li, W. V. Kalina, D. Cooper, R. W. Frenck, Jr., L. L. Hammitt, O. Tureci, H. Nell, A. Schaefer, S. Unal, D. B. Tresnan, S. Mather, P. R. Dormitzer, U. Sahin, K. U. Jansen, W. C. Gruber, C. C. T. Group, Safety and Efficacy of the BNT162b2 mRNA Covid-19 Vaccine. N Engl J Med 383, 2603–2615 (2020).

61. L. R. Baden, H. M. El Sahly, B. Essink, K. Kotloff, S. Frey, R. Novak, D. Diemert, S. A. Spector, N. Rouphael, C. B. Creech, J. McGettigan, S. Khetan, N. Segall, J. Solis, A. Brosz, C. Fierro, H. Schwartz, K. Neuzil, L. Corey, P. Gilbert, H. Janes, D. Follmann, M. Marovich, J. Mascola, L. Polakowski, J. Ledgerwood, B. S. Graham, H. Bennett, R. Pajon, C. Knightly, B. Leav, W. Deng, H. Zhou, S. Han, M. Ivarsson, J. Miller, T. Zaks, C. S. Group, Efficacy and Safety of the mRNA-1273 SARS-CoV-2 Vaccine. N Engl J Med 384, 403–416 (2021).

62. E. J. Haas, F. J. Angulo, J. M. McLaughlin, E. Anis, S. R. Singer, F. Khan, N. Brooks, M. Smaja, G. Mircus, K. Pan, J. Southern, D. L. Swerdlow, L. Jodar, Y. Levy, S. Alroy-Preis, Impact and effectiveness of mRNA BNT162b2 vaccine against SARS-CoV-2 infections and COVID-19 cases, hospitalisations, and deaths following a nationwide vaccination campaign in Israel: an observational study using national surveillance data. The Lancet 397, 1819–1829 (2021).

63. UK Heath Security Agency, COVID-19 vaccine surveillance report (Week 17, 28 April 2022). https://assets.publishing.service.gov.uk/government/uploads/system/uploads/attachment_data/file/1072064/Vaccine-surveillance-report-week-17.pdf

64. F. C. M. Kirsebom, N. Andrews, J. Stowe, S. Toffa, R. Sachdeva, E. Gallagher, N. Groves, A.-M. O’Connell, M. Chand, M. Ramsay, J. L. Bernal, COVID-19 vaccine effectiveness against the omicron (BA.2) variant in England. The Lancet Infectious Diseases.

65. J. L. Anderson, An ensemble adjustment Kalman filter for data assimilation. Mon. Weather Rev. 129, 2884–2903 (2001).

